# Single-gene transcripts for subclinical TB: an individual participant data meta-analysis

**DOI:** 10.1101/2024.07.04.24309943

**Authors:** James Greenan-Barrett, Simon C. Mendelsohn, Thomas J. Scriba, Mahdad Noursadeghi, Rishi K. Gupta

**Affiliations:** UCL Respiratory, Division of Medicine, University College London, London, United Kingdom; South African Tuberculosis Vaccine Initiative, Division of Immunology, Department of Pathology and Institute of Infectious Disease and Molecular Medicine, University of Cape Town, Cape Town, South Africa; UCL Division of Infection and Immunity, University College London, London, United Kingdom

## Abstract

**Background:** Translation of blood RNA signatures may be accelerated by identifying more parsimonious biomarkers. We tested the hypothesis that single-gene transcripts provide comparable accuracy for detection of subclinical TB to multi-gene signatures and benchmarked their clinical utility to interferon-y release assays (IGRAs).

**Methods:** We identified datasets where participants underwent RNA sampling and at least 12 months of follow-up for progression to TB. We performed a one-stage individual participant data meta-analysis to compare multi-gene signatures against single-gene transcripts to detect subclinical TB, defined as asymptomatic prevalent or incident TB (diagnosed ≥21 days from enrolment, irrespective of symptoms) over a 12-month interval. We performed decision curve analysis to evaluate the net benefit of using RNA biomarkers and IGRA, alone or in combination, compared to treating all or no individuals with preventative treatment.

**Results:** We evaluated 80 single-genes and eight multi-gene signatures in a pooled analysis of four RNAseq and three qPCR datasets, comprising 6544 total samples and including 283 samples from 214 individuals with subclinical TB. Five single-gene transcripts were equivalent to the best-performing multi-gene signature over 12 months, with areas under the receiver operating characteristic curves ranging from 0.75-0.77, but none met the WHO minimum target product profile (TPP) for a TB progression test. IGRA demonstrated much lower specificity in higher burden settings, while sensitivity and specificity of RNA biomarkers were consistent across settings. In higher burden settings, RNA biomarkers had greater net benefit than IGRA, which offered little clinical utility over treating all with preventative therapy. In low burden settings, IGRA approximated the TPP and offered greater clinical utility than RNA biomarkers, but combining both tests provided the highest net benefit for services aiming to treat <50 people to prevent a single case.

**Interpretation:** Single-gene transcripts are equivalent to multi-gene signatures for detection of subclinical TB, with consistent performance across settings. Single transcripts demonstrate potential clinical utility to stratify treatment, particularly when used in combination with IGRA in low burden settings.

## Research in Context

### Evidence before this study

With increasing recognition of the spectrum of tuberculosis and the importance of subclinical TB, the International Consensus for Early TB (ICE-TB) group recently developed a framework to classify disease states and called for diagnostic development to detect the subclinical states. We performed a systematic search of PubMed from inception to June 10, 2024 using terms for ‘tuberculosis’, ‘subclinical’ and ‘RNA’, without language restrictions. Multiple studies have discovered or validated multi-gene blood RNA signatures for TB, including to discriminate individuals who progress to clinical TB from non-progressors. In our previous head-to-head evaluation of these signatures, we demonstrated that eight signatures had equivalent performance to predict progression to TB. More parsimonious biomarkers, such as single-gene transcripts, may facilitate clinical translation. However, no previous studies have systematically compared the performance of single-gene transcripts to multi-gene signatures. Moreover, the clinical utility of RNA biomarkers to detect subclinical TB and guide treatment decisions, compared to alternative strategies including using interferon-gamma release assays (IGRAs), remains untested.

### Added value of this study

To our knowledge, this is the largest pooled analysis of RNA biomarkers to predict progression to clinical TB, the first head-to-head comparison of single-gene transcripts to multi-gene signatures and the first RNA analysis to align with the ICE-TB definitions. We tested 80 single-genes and eight multi-gene signatures to detect subclinical TB in a pooled dataset from four RNAseq and three qPCR datasets, comprising over 6500 RNA samples. We show that five co-correlated single-gene transcripts were equivalent to the best-performing multi-gene signature to detect subclinical TB over a 12-month interval, but none met the WHO minimum target product profile (TPP) for a TB progression test. Discriminative performance of single-gene transcripts waned over increasing time intervals from sampling. BATF2, the transcript with the highest discrimination point estimate, showed consistent performance across settings. In contrast, IGRA performance was heterogenous, approximating the WHO minimum TPP in low TB burden settings, but demonstrating poor specificity in higher burden settings. Decision curve analysis comparing the clinical utility of BATF2 and IGRA showed that BATF2 offered highest net benefit in high burden settings. In low burden settings, IGRA offered greater net benefit, while a two-step approach combining both tests achieved the greatest net benefit for services aiming to treat fewer than 50 people to prevent a TB case.

### Implications of all the available evidence

Single-gene transcripts perform as well as multi-gene signatures for subclinical TB. These findings may simplify RNA biomarker testing, encourage commercial competition and facilitate translation of this technology to clinical practice. RNA biomarkers demonstrate clinical utility to direct treatment decisions, as a stand-alone test in high burden settings and in combination with IGRA in low burden settings. Further interventional studies are required to evaluate the clinical and cost-effectiveness of serial RNA biomarker testing to stratify delivery of preventative therapy, for example among high-risk contacts in high burden countries, or using a two-step testing approach in combination with IGRA in low burden settings.

## Introduction

Despite global efforts, tuberculosis (TB) remains a leading cause of morbidity and mortality worldwide, causing 10.6 million cases and 1.3 million deaths in 2022, with a disproportionate burden on disadvantaged communities^1,2^. In response to increased recognition of the spectrum of TB, the International Consensus for Early TB (ICE-TB) group recently developed a framework to classify disease states. The ICE-TB framework divides TB disease into subclinical or clinical based on signs and symptoms, with further subdivisions into infectious and non-infectious based on detection of aerosolised or expectorated *M tuberculosis*^3^. Targeting the subclinical disease state, potentially with truncated treatment regimens^4^, may prevent progression to clinical disease and reduce risk of onward transmission.

Current prognostic tests for TB, such as the tuberculin skin test (TST) and interferon-γ release assay (IGRA), have low positive predictive values (PPVs) for progression to clinical disease^5–7^, resulting in unnecessary TB preventative therapy (TPT) for most individuals. This is financially burdensome in healthcare systems and adverse effects are estimated to affect 3.7% of TPT-treated individuals^8^. Measurement of blood RNA levels can be used to detect changes in host gene expression in response to TB disease. Multiple RNA signatures have been discovered that have promising diagnostic accuracy for clinical TB^9–20^ or to predict progression to clinical disease^21–24^. In a previous analysis comparing the performance of 17 RNA signatures to predict progression to clinical disease, we demonstrated that eight signatures performed equivalently, were co-correlated and shared common upstream pathways^25^.

Development of near-patient, cartridge-based prototype platforms, such as the Cepheid MTB-Host Response (MTB-HR) assay have paved the way for clinical translation of blood RNA signatures. However, this progress may be further accelerated by further simplification of multi-gene signatures to single-gene biomarkers. In view of the co-correlation and common regulators of the genes that comprise the most accurate signatures to date, multiple genes may not offer orthogonal value. In this study, we used a pooled dataset of studies with blood RNA sampling and longitudinal follow-up for TB to hypothesise that single-gene transcripts will perform as well as multi-gene signatures for detection of subclinical TB. We also sought to benchmark the diagnostic performance and clinical utility of RNA biomarkers to IGRA, stratified by TB burden.

## Methods

### Data sources and preparation

We performed a systematic search to identify datasets where participants underwent whole blood RNA sampling with at least 12 months of follow-up for development of clinical TB (supplementary methods). Studies using genome-wide (RNAseq or microarray) or targeted transcriptional profiling (qPCR or NanoString quantification) were included. We included four RNAseq datasets from our previous individual participant data meta-analysis (IPD-MA)^25^ and three subsequent studies using qPCR^26–28^. All included RNA datasets were publicly available.

The four previously included RNAseq datasets were mapped, batch corrected, and integrated into a single pooled dataset using transcripts per million measurements, as previously described^25^. Our data preparation pipeline for qPCR studies is described in the supplementary methods. We included 80 single-genes that were present in the RNAseq dataset and at least one qPCR study (CORTIS-01/HR^26,27^ or REPORT-Brazil^28^). We calculated scores for eight existing RNA signatures that were included in our previous analysis (supplementary methods)^29^. We standardised signatures and single-gene transcripts within each RNAseq and qPCR dataset by converting to z-scores (supplementary methods), before combining the z-score transformed datasets into a pooled dataset.

### Outcome definitions

We used the original study definitions as a reference standard for TB in each cohort. We sought to align our terminology with the ICE-TB consensus classification^3^. Clinical TB was defined as symptomatic prevalent TB cases. Subclinical TB was defined as asymptomatic prevalent TB cases (approximates to ‘subclinical TB, infectious’) or incident TB cases (approximates to ‘subclinical TB, non-infectious’). The ‘subclinical, non-infectious’ state is based on the presence of “macroscopic pathology”, for which there is no gold standard. We therefore used progression to incident TB as a reference standard to approximate this, based on the assumption that macroscopic pathology would have been detectable at the time of blood RNA sampling if investigated with sufficient resolution. Prevalent TB was defined as a TB diagnosis made less than 21 days after RNA sampling whereas incident TB was defined as a TB diagnosis ≥21 days. Non-progressors were defined as individuals who remained TB-free during follow-up.

### Analysis

All analyses were done using R (version 4.4.0). We performed a one-stage IPD-MA to calculate the accuracy of candidate signatures and transcripts to discriminate subclinical TB from non-progressors, stratified by interval from sampling to disease. Since our initial analyses demonstrated similar accuracy of RNA biomarkers for each study, our primary analysis assumed common accuracy across studies, as previously^25^. The primary analysis was over a 12-month interval from sampling, with further stratified analyses over 0-3, 0-6, 0-15, 6-12, and 12-15 months. For the primary analysis, non-progressor samples with less than 12 months of follow-up from sampling were excluded. We excluded individuals who received TPT from the primary analysis since this affects progression risk and may be differential among those with higher, compared to lower, RNA biomarker scores. Where datasets included serial samples from the same individuals, we considered serial samples as independent since intra-individual variance was similar to inter-individual variance (supplementary figure 7). Where candidate signatures were originally derived from included datasets, we excluded these datasets when evaluating the accuracy of that signature.

Accuracy of candidate signatures and transcripts was quantified by the area under the receiver operating curve (AUROC) with 95% confidence intervals (CIs). Sensitivity and specificity were calculated at the maximum Youden index, giving equal weighting to sensitivity and specificity, and benchmarked against the WHO minimum Target Product Profile (TPP) parameters for predicting progression to TB over 2 years (≥75% sensitivity and ≥75% specificity)^04^/07/2024 14:57:00. AUROCs of single-gene transcripts were compared to the best performing multi-gene signature using the pairwise Delong test, with multiple testing correction using the Benjamini-Hochberg approach. Signatures and transcripts with adjusted p values >0.05 were considered equivalent. Correlation of equivalent transcripts was assessed using Spearman rank correlation. We also evaluated expression and AUROCs of the best-performing single gene transcript across different disease states; clinical TB, ‘subclinical TB, infectious’, and ‘subclinical TB, non-infectious’.

We then compared the diagnostic performance and clinical utility of the single-gene transcript with the highest AUROC point estimate to IGRA, in a head-to-head analysis among participants for whom results of both tests were available, over a 12-month interval from sampling. These analyses were stratified by setting, defined as low TB burden where incidence was <50 per 100,000, and high TB burden above this. Among individuals with serial IGRA samples, low intra-individual variance indicated high correlation between serial samples (supplementary figure 7); we therefore included only one sample per individual, sampled at random. We used thresholds of the maximum Youden index for the transcript and the standard cut-off of 0.35 IU/ml for the QuantiFERON-TB assay. We also explored an approach using a combination of the transcript and IGRA, where only those positive for both tests are offered treatment. We compared the sensitivity, specificity and PPV of these approaches based on a 1% and 2% prior probability, with the latter based on 1-year incidence rates in high-risk close contacts^30,31^.

To evaluate the clinical utility of the RNA biomarker, IGRA and combined testing approaches, we performed decision curve analysis. Decision curve analysis quantifies the trade-off between correctly identifying true TB progressors and incorrectly identifying false-positives, as ‘net benefit’^32^. Net benefit is calculated across a range of weightings for the false-positives, defined as the threshold probability. Threshold probability is the risk of disease at which a clinician or patient would opt for an intervention such as treatment and relates to the number-willing-to-treat to prevent a single case of disease. We calculated net benefit using the best performing transcript or IGRA to guide preventative treatment compared to the default strategies of treating all individuals or no individuals, across a range of threshold probabilities. Since the contributing datasets included case-control analyses, we fixed the cumulative TB risk as 1% and 2% in our decision curve analyses.

Finally, we estimated the number-needed-to-treat with preventative therapy to prevent a single TB case for these testing approaches compared to a default strategy of treating all individuals. We assumed a constant treatment effect of 80% and fixed the cumulative TB risk as 1% and 2%.

### Sensitivity analyses

We performed four sensitivity analyses. Firstly, we performed a two-stage IPD-MA where we calculated accuracy of signatures and transcripts for each contributing cohort, explored between study heterogeneity, and performed a random-effects meta-analysis to calculate pooled AUROCs (using the *metafor* package in R^33^). Secondly, we included participants commencing TPT in the analysis. Thirdly, we included only one sample per individual, sampled at random. Finally, we included datasets from which signatures were originally derived in the accuracy calculation for that signature.

## Results

### Overview of contributing studies

Four RNAseq datasets and three qPCR datasets were included (table 1). The RNAseq datasets included the Adolescent Cohort Study (ACS) of IGRA/TST positive individuals from South Africa^21^, the Grand Challenges 6-74 (GC6-74) study of household contacts of TB patients from South Africa, The Gambia, and Ethiopia^22^, and two UK close contact studies from London^24^ and Leicester^34^. The qPCR datasets included the Correlate of Risk Targeted Intervention Study (CORTIS-01) study of healthy volunteers from TB endemic communities in South Africa^26^, the CORTIS-HR study consisting of people living with HIV (PLHIV) from TB endemic communities in South Africa^27^, and the Regional Prospective Observational Research for Tuberculosis Brazil (REPORT-Brazil) study of close contacts of TB patients from Brazil^28^.

**Table 1:** Characteristics of studies included in meta-analysis.

|  | ACS | CORTIS-01 | CORTIS-HR | GC6-74 | Leicester Contacts | London Contacts | REPORT-Brazil |
| --- | --- | --- | --- | --- | --- | --- | --- |
| **Method of RNA profiling** | RNAseq | RT-qPCR | RT-qPCR | RNAseq | RNAseq | RNAseq | RT-qPCR |
| **Study design** | Nested case-control | Randomised controlled trial* | Cohort | Nested case-control | Cohort | Cohort | Cohort |
| **Population** | Positive IGRA/TST | Endemic community | PLHIV in endemic community | Household contacts | Close contacts | Close contacts | Close contacts |
| **Age** | 12-18 | ≥18 | ≥18 | 10-60 | ≥16 | ≥18 | ≥18 |
| **TB burden of setting** | High burden | High burden | High burden | High burden | Low burden | Low burden | Low burden |
| **Country** | South Africa | South Africa | South Africa | South Africa, The Gambia, Ethiopia | UK | UK | Brazil |
| **Baseline screening** | Clinical evaluation | 2x sputum | 2x sputum | Clinical evaluation | Clinical evaluation + CXR | Clinical evaluation + CXR | Clinical evaluation |
| **Study case definition** | 2x positive smear or 1x positive culture | 2x positive Xpert MTB/RIF or Xpert Ultra or culture | 2x positive Xpert MTB/RIF or Xpert Ultra or culture | Positive culture or clinically diagnosed | Positive culture or Xpert MTB/RIF | Positive culture or clinically diagnosed | Positive culture or clinically diagnosed |
| **Duration of follow-up (months)** | 24 | 15 | 15 | 24 | 24 | 22.8 | 24 |
| **RNA sample timing** | Baseline & 6/12/18/24 months | Baseline | Baseline | Baseline & 6/18 months | Baseline | Baseline | Baseline & 6 months |
| **Total participants** | 144 | 2496 | 404 | 334 | 104 | 324 | 1379 |
| **Total RNA samples** | 318 | 2496 | 404 | 412 | 104 | 324 | 2472 |

\*The cohort was enriched for RISK11-positive individuals. The RISK11-positive individuals were randomised to TPT or no TPT. The RISK11-negative individuals did not receive TPT.
Legend: CXR, chest x-ray; PLHIV, people living with HIV

ACS and GC6-74 studies were nested case-control studies within larger prospective cohort studies. CORTIS-01 was a randomised control trial where a cohort of healthy participants first underwent measurement of the RISK11 signature (Darboe11). RISK11-positive individuals were randomised to either receive TPT or not, whilst the RISK11-negative individuals did not receive TPT. Only a subset of randomly sampled RISK11-negative participants were included to enrich the study cohort for RISK11-positive participants. All other studies were observational cohort studies. All studies performed RNA sampling at baseline; ACS, GC6-74, and REPORT-Brazil also performed serial, 6-monthly sampling. REPORT-Brazil, Leicester contacts, and London contacts were from countries with a low TB burden, whereas the remainder were from countries with a high TB burden. CORTIS-01 and CORTIS-HR performed 15 months of follow-up whereas the remaining studies performed 22.8-24 months of follow-up. TB case definitions varied by study; all included microbiologically confirmed cases (culture +/-smear/PCR) while GC6-74, London contacts, and REPORT-Brazil also included clinically diagnosed cases.

### Overview of included samples

In total, 6530 samples from 5185 individuals were included in the primary analysis, with a total of 283 subclinical TB samples – 39 asymptomatic prevalent samples and 244 incident samples (supplementary figure 8). Baseline characteristics and risk of TB disease for each study are shown in supplementary table 1.

Three of the included eight signatures were derived from included datasets – Darboe11 and Penn-Nicholson6 from ACS, and Suliman4 from GC6-74 – so the original datasets were excluded from the evaluation of signature performance in the primary analysis. Distributions of gene z-scores after standardization were similar between datasets and there was little heterogeneity in AUROCs of transcripts between datasets (supplementary figures 4B & 10). We therefore proceeded to a one-stage IPD-MA as the primary analysis.

### Accuracy of signatures and single transcripts for subclinical TB

The multi-gene signature with the highest AUROC for discrimination of subclinical TB from non-progressors over 12 months from sampling was Roe3 (AUC 0.77 [95% CI 0.73-0.81]), which was then used for pairwise comparison with the single-gene transcripts. Five single-gene transcripts had equivalent AUROCs to Roe3; BATF2 (0.77 [0.73-0.81]), FCGR1A/B (0.77 [0.73-0.81]), ANKRD22 (0.77 [0.72-0.81]), GBP2 (0.75 [0.71-0.79]), and SERPING1 (0.75 [0.71-0.79]) (table 2). These transcripts demonstrated moderate-strong correlation (0.54-0.81) using Spearman rank (supplementary figure 9). Using maximum Youden index thresholds, none of the transcripts met the WHO minimum TPP parameters for progression tests over 12 months. Discriminative performance of single-gene transcripts diminished gradually over increasing time intervals from sampling to disease, with poor discrimination over 12-15 months (supplementary table 2). BATF2 expression and discriminative performance compared to non-progressors across different disease states is shown in figure 1. Notably, discriminative performance of BAFT2 was highest for clinical TB (AUROC 0.93 [0.87-0.99]).

**Table 2:**
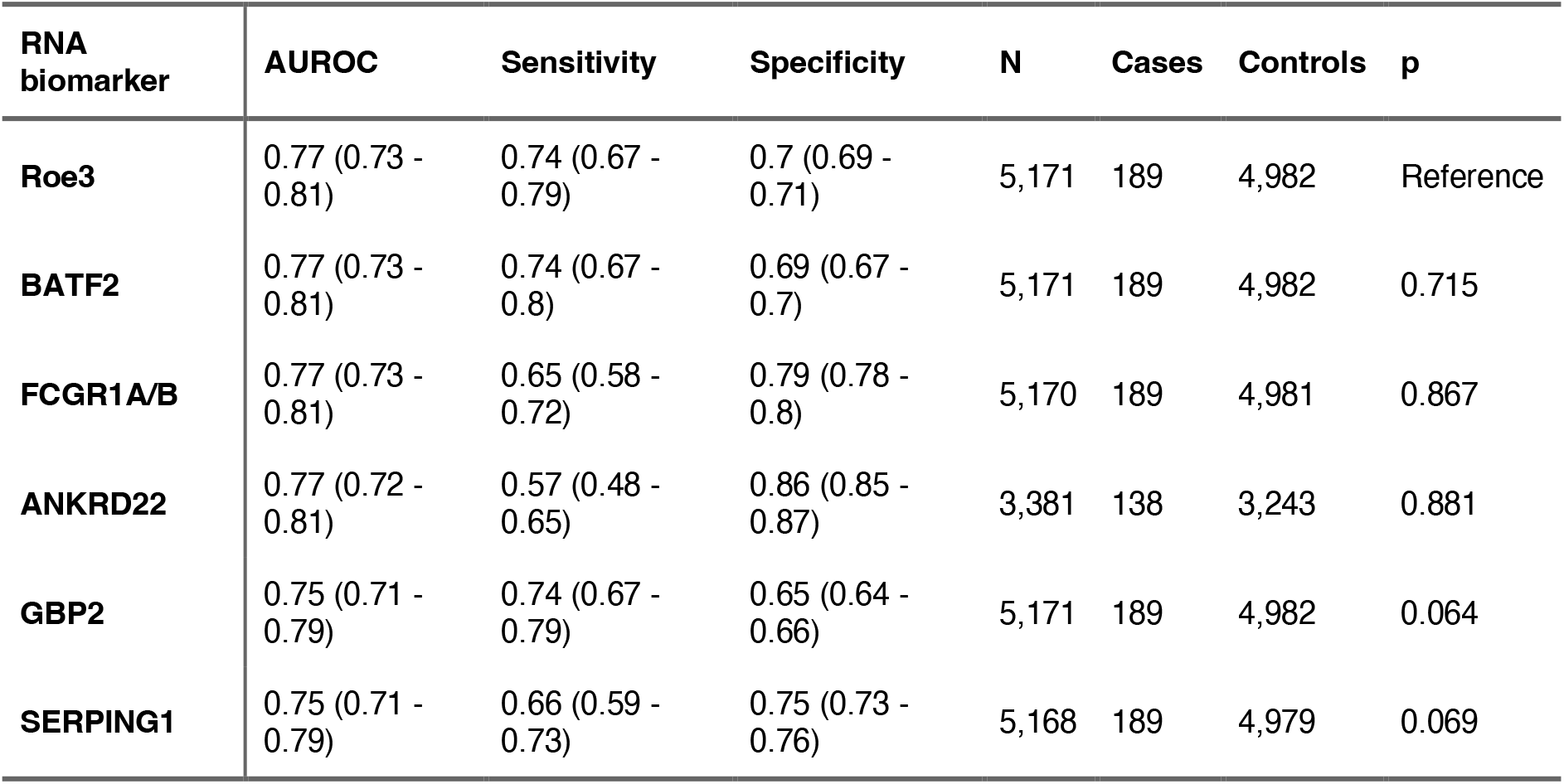
Performance metrics of equivalent single-gene transcripts for subclinical TB. Performance metrics of single-gene transcripts with equivalent performance to the best performing multi-gene signature (Roe3) to discriminate subclinical TB cases from non-progressors at an interval of 12 months from sampling to disease are shown. Equivalence to Roe3 was defined as an adjusted p value >0.05 in the pairwise Delong test. Performance metrics include estimates and 95% confidence intervals for the Area Under the Receiver Operating Curve (AUROC) as well as sensitivity and specificity at the maximum Youden index calculated from the one-stage meta-analysis.

| RNA biomarker | AUROC | Sensitivity | Specificity | N | Cases | Controls | p |
| --- | --- | --- | --- | --- | --- | --- | --- |
| **Roe3** | 0.77 (0.73 - 0.81) | 0.74 (0.67 - 0.79) | 0.7 (0.69 - 0.71) | 5,171 | 189 | 4,982 | Reference |
| **BATF2** | 0.77 (0.73 - 0.81) | 0.74 (0.67 - 0.8) | 0.69 (0.67 - 0.7) | 5,171 | 189 | 4,982 | 0.715 |
| **FCGR1A/B** | 0.77 (0.73 - 0.81) | 0.65 (0.58 - 0.72) | 0.79 (0.78 - 0.8) | 5,170 | 189 | 4,981 | 0.867 |
| **ANKRD22** | 0.77 (0.72 - 0.81) | 0.57 (0.48 - 0.65) | 0.86 (0.85 - 0.87) | 3,381 | 138 | 3,243 | 0.881 |
| **GBP2** | 0.75 (0.71 - 0.79) | 0.74 (0.67 - 0.79) | 0.65 (0.64 - 0.66) | 5,171 | 189 | 4,982 | 0.064 |
| **SERPING1** | 0.75 (0.71 - 0.79) | 0.66 (0.59 - 0.73) | 0.75 (0.73 - 0.76) | 5,168 | 189 | 4,979 | 0.069 |

**Figure 1:**
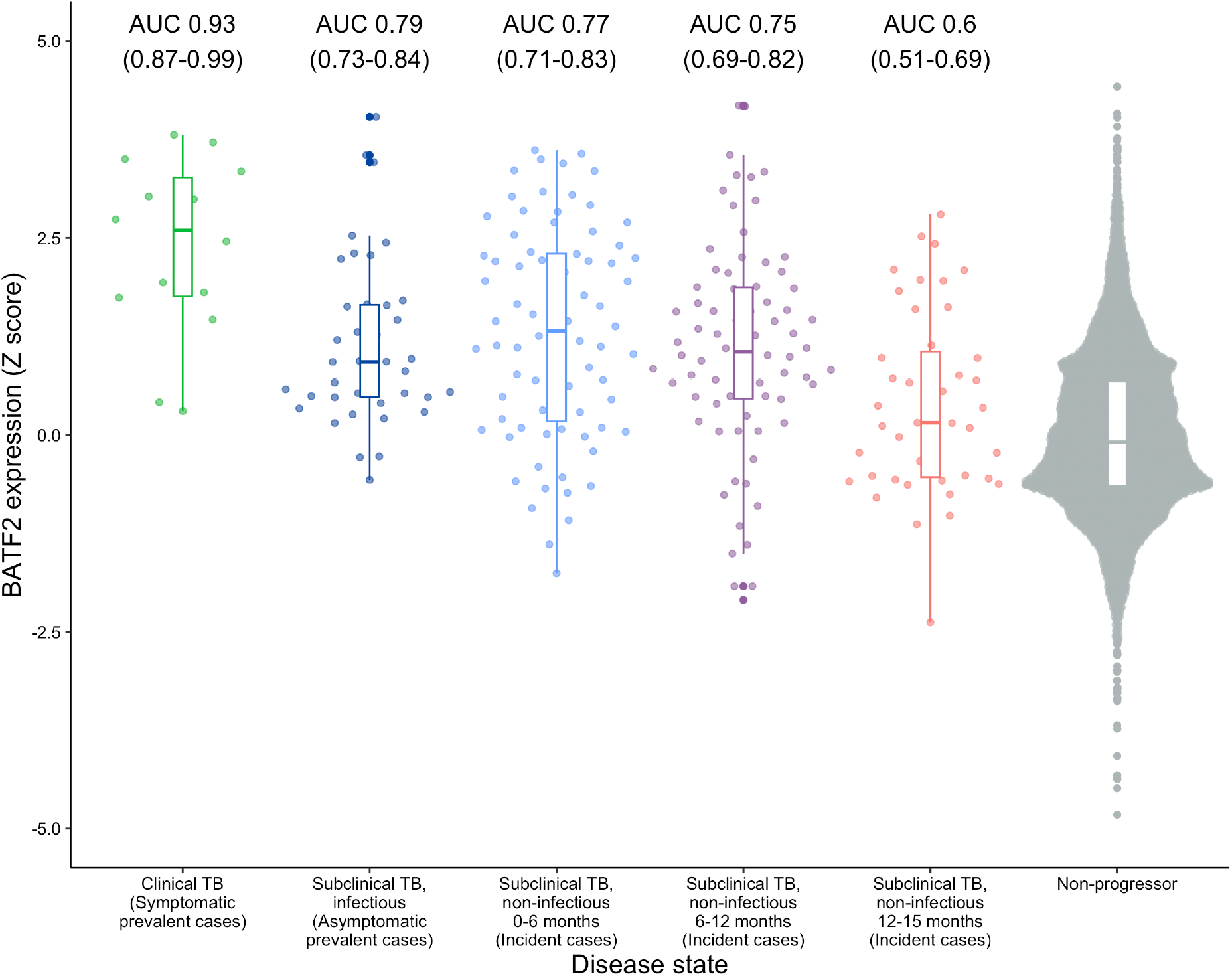
BATF2 expression and diagnostic accuracy for different disease states. Expression (Z-score transformed) of the best performing single-gene transcript, BATF2, is shown as a combined boxplot and scatterplot, stratified by disease state. Disease state is classified according to ICE-TB consensus definitions and descriptive terms are also shown. Boxes represent interquartile range and median values. Grey dots represent individual samples and black dots represent outliers. Area Under the Curve (AUC) with 95% confidence intervals of BATF2 to discriminate the disease state from non-progressors is also shown.

### Comparison of accuracy to IGRA

Comparison of the diagnostic performance of BATF2, IGRA, and a combined approach of BATF2 and IGRA, stratified by setting and benchmarked against the WHO minimum TPP performance criteria (figure 2A) showed that whilst the sensitivity of IGRA was similar across settings (87-88%), specificity was markedly lower in high burden settings (32% [95% CI 30-35%]) compared to low burden settings (74% [72-76%]) resulting in PPVs of 1.3% (1.1-1.4%) and 3.3% (2.4-3.9%) respectively (supplementary table 4). In contrast, BATF2 performance was consistent across settings, with PPVs of 2.3% (1.8-2.7%) and 2.4% (1.6-3.1%) in high and low burden settings respectively. In high burden settings, the combined approach resulted in a slight increase in specificity compared to BATF2 alone (77% [54-76%] versus 67% [64-69%]) but similar PPVs (2.8% [2.1-3.5%] versus 2.3% [1.8-2.7%]). In contrast, a large increase in specificity (92% [CI 90-93%] versus 72% [70-74%]) and PPV (6.8% [3.8-9.9%] versus 2.4% [1.6-3.1%]) was achieved in the low burden settings when using the combined approach compared to BATF2 alone, albeit with some loss of sensitivity (58% [39-76%] versus 67% [47-82%]). None of the testing approaches met WHO minimum TPP parameters over 12 months, although IGRA accuracy approached the benchmarks in the low burden settings. Using a 2% prior probability, PPVs approximately doubled without changing the overall pattern of results (supplementary figure 12).

**Figure 2:**
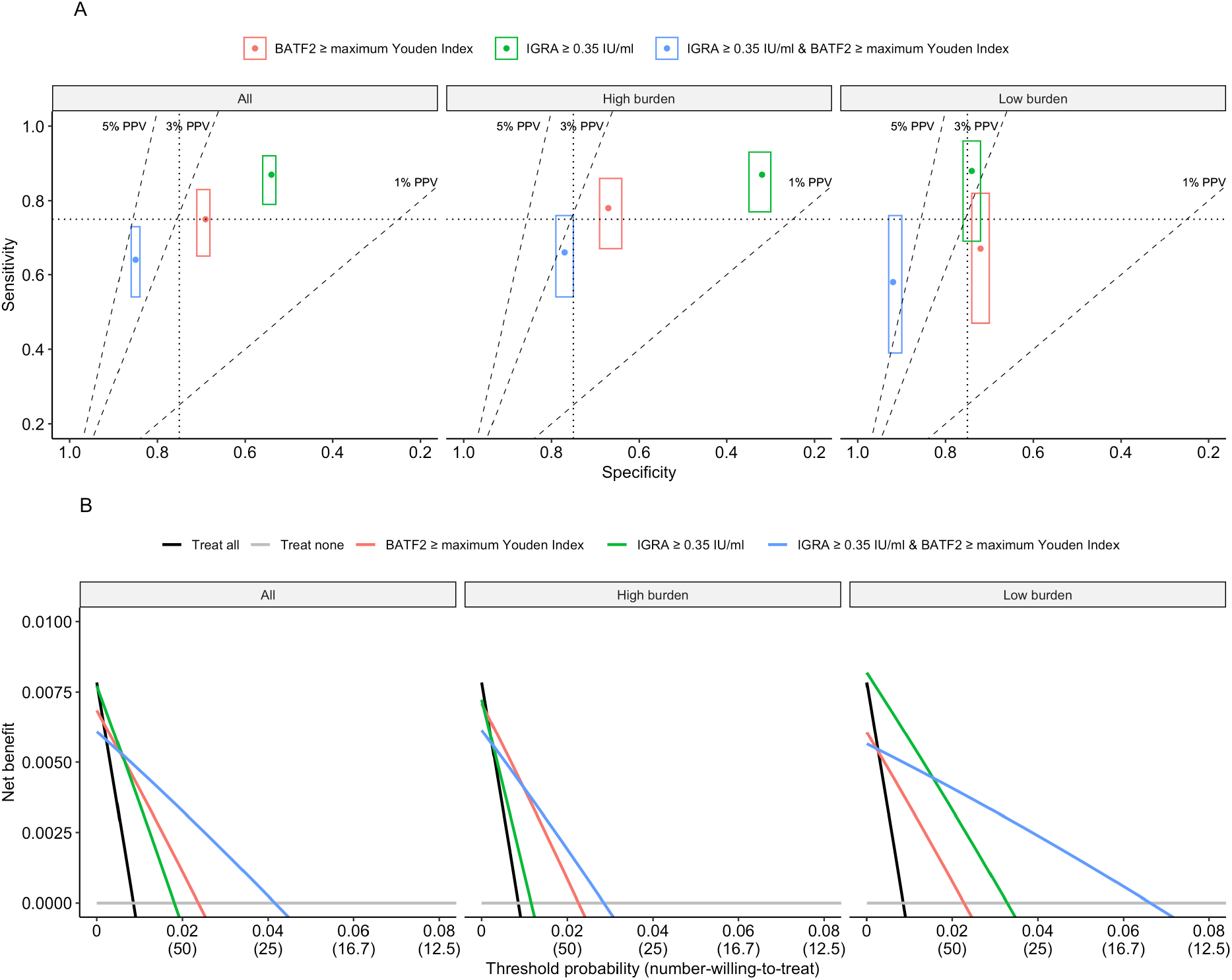
Diagnostic performance for subclinical TB of BATF2, IGRA and a combined approach shown in receiver operating space and in a decision curve analysis, by setting. Figure 2A compares the diagnostic performance for subclinical TB over a 0-12 month interval of BATF2 (threshold set at maximum Youden Index), IGRA (threshold set at standard cutoff of 0.35 IU/ml) and a combined approach of BATF2 and IGRA in the receiver operating space with sensitivity on the y axis and 1-specificity on the x axis, stratified by setting. Only participants with results for both tests were included in this analysis. Shown as point estimates with 95% confidence intervals (boxes). Dotted lines represent the 75% WHO minimum Target Product Profile sensitivity and specificity for a TB progression test. Dashed lines represent positive predictive values of 1%, 3% and 5%, based on a 1% prior probability. Underlying data are shown in supplementary table 4. Figure 2B is a decision curve analysis where each test is compared to default strategies of treating all or treating no persons, stratified by setting. Threshold probability is the risk of TB disease at which a clinician or patient would opt for preventative therapy and is the reciprocal of the number-willing-to-treat to prevent a single case. Net benefit is calculated at a range of threshold probabilities as the true positive rate minus a weighted false positive rate, where the weighting is the threshold probability. Since the contributing datasets included case-control analyses, the cumulative TB risk was fixed at 1%.

### Clinical utility analysis

Decision curve analysis of these testing approaches varied by setting (figure 2B). In high burden settings, IGRA offered minimal additional net benefit over a treat all strategy. BATF2 offered greater net benefit than IGRA and, using prior probability of 1%, had the highest net benefit across threshold probabilities between 0.4% and 2.2%, equating to a number-willing-to-treat to prevent a single case (NWT) of 45-250. Above this threshold, where NWT was less than 45, treating none was best. The combined approach (where only those positive for both tests are offered treatment) achieved only slightly greater net benefit than BATF2 alone at threshold probabilities of 1-3%, equating to a NWT of 33-100. In contrast, in low burden settings, IGRA outperformed BATF2 across all threshold probabilities. IGRA offered the highest net benefit at threshold probabilities under 2%, equating to a NWT of over 50, whereas the combined approach offered the highest net benefit at threshold probabilities of 2-7%, equating to a NWT of 14-50. Using a higher prior probability of 2%, findings were similar, but net benefit for all strategies was shifted to the right on the threshold probability scale. For example, in higher burden settings, BATF2 offered greater net benefit over IGRA and a treat all strategy at threshold probabilities of 0.7-4.5%, equating to a NWT 22-143 (supplementary figure 12).

Number-needed-to-treat (NNT) estimates to prevent a single TB case for the testing approaches, compared to a treat all strategy, are shown in figure 3A. In high burden settings, performing IGRA testing resulted in slightly lower NNT estimates than treating all (98 [88-114] versus 125). Compared to IGRA, NNTs were significantly lower using BATF2 (54 [46-68]) or a combined approach (44 [34-59]). In low burden settings, NNTs of IGRA and BATF2 were similar (38 [32-51] and 53 [40-80] respectively). Using the combined approach resulted in a lower NNT of 18 (13-33). Due to superior specificity, lower NNT estimates were achieved by IGRA (either alone or in combination with BATF2) in low burden versus high burden settings. Using a 2% prior probability roughly halved the NNT with a similar pattern (figure 3B).

**Figure 3:**
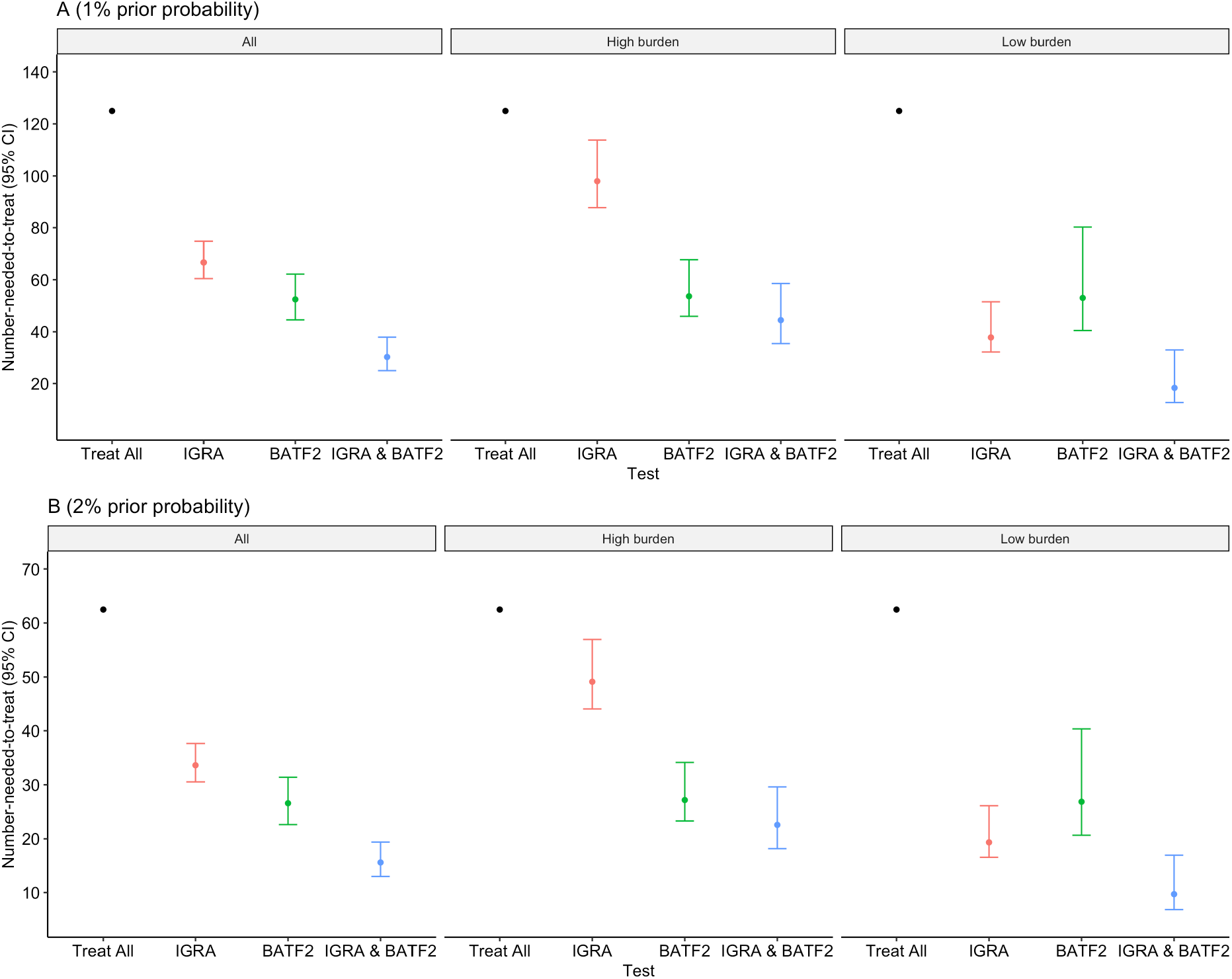
Number-needed-to-treat to prevent a single TB case using different testing strategies, by setting, using 1% and 2% prior probabilities. Estimated number-needed-to-treat (NNT) with preventative therapy to prevent a single TB case is shown for the default strategy of treating all compared to test stratified treatment using BATF2 (threshold set at maximum Youden Index), IGRA (threshold set at standard cutoff of 0.35 IU/ml) and a combined approach of BATF2 and IGRA. Shown as point estimates and 95% confidence intervals (bars). An estimated treatment effect of 80% was used. Since the contributing datasets included case-control analyses, the cumulative TB risk was fixed at 1% (A) and 2% (B).

### Sensitivity analyses

A two-stage meta-analysis to calculate pooled AUROCs resulted in similar findings to the primary analysis (supplementary table 5). Similarly, we observed similar AUROCs when including recipients of TPT (excluding the Darboe-11 positive participants randomised to systematically receive TPT in CORTIS-01), including only one RNA sample per individual, or when including datasets from which signatures were originally derived in the accuracy calculation for that signature (supplementary table 6).

## Discussion

To the best of our knowledge, we report the largest pooled RNA biomarker analysis in subclinical TB to date, including over 6500 RNA samples, and the first comprehensive head-to-head analysis comparing single-gene transcripts to multi-gene signatures. This is also the first evaluation of RNA biomarkers to align with the ICE-TB classification. We showed that five single-gene transcripts perform equivalently to the best multi-gene signature for discrimination of subclinical TB from non-progressors over 12 months. RNA biomarker performance was consistent across settings. In contrast, IGRA performance varied markedly, with poor performance in higher burden settings. In decision curve analysis we demonstrate that, in high burden settings, RNA biomarker testing offers clinical utility over IGRA, which has minimal benefit over treating all with preventative therapy. In low burden settings, IGRA was the best single test and approximated the WHO TPP, with greater net benefit than RNA biomarker testing. However, for services aiming to treat fewer than 50 people to prevent a TB case, a two-step combined testing approach improves specificity and is superior.

Development of the Cepheid MTB-HR prototype has demonstrated that translation to a near-patient platform is feasible. Whilst the cost of such platforms is currently unknown, it is unlikely to exceed the WHO TPP maximum of $100 per test, based on the cost of an IGRA^35^. Our findings may facilitate translation of RNA biomarker technology to clinical practice by encouraging commercial competition using measurement of any one of best performing single genes at lower cost. Nonetheless, none of the transcripts met the WHO TPP minimum sensitivity and specificity, even over a 12-month interval, although this was almost achieved by IGRA in the low burden setting (74% specificity and 88% sensitivity). In the high burden setting, the WHO TPP seem an unrealistic aim that is unlikely to be achieved over two years with a biomarker targeting early disease, although serial testing may improve overall performance. Combining different modalities of tests may improve specificity, albeit at greater cost. A universal testing strategy may be challenging to achieve if test performance is heterogenous across settings; rather tailored strategies may be required based on TB burden. We demonstrate this by showing that in low burden settings IGRA remains a useful test; a combined approach using IGRA and blood RNA biomarkers shows additional promise and warrants further evaluation. However, in high burden settings, the high prevalence of *Mtb* sensitisation means that IGRA has poor specificity and thus minimal utility. Greater specificity may be achieved with better measures of recent *Mtb* infection, for example *Mtb*-specific T cell activation^36^. Alternatively, combining molecular approaches with radiological testing, such as digital chest radiographs^37^, as a method to detect macroscopic pathology, may also improve performance.

Our findings may provide some insights into host immune responses in early TB. The co-correlation of best performing single-gene transcripts is consistent with previous findings of shared upstream interferon and tumour necrosis factor signalling pathways^25^. This explains why a single transcript is a sufficient measure of this immune response and combining these transcripts into multigene signatures does not offer orthogonal value. The consistent performance of RNA biomarkers across settings suggests that this a common host response across populations. Likewise, the similar performance in CORTIS-HR suggests that this pathway is preserved in PLHIV, though previous data have demonstrated likely upregulation of common type-I interferon responses in untreated HIV^38^. The fall in discriminative performance after 12 months may be reflective of de novo infection following signature measurement in high burden settings, or that there is a minimal host response in earlier subclinical disease. The equivalent performance of multiple co-correlated RNA biomarkers, which has also been reported previously^25,27,38^, suggests that future discovery and validation of signatures using similar approaches is unlikely to yield a test with better performance. To date, discovery approaches have largely focused on identifying differentially expressed transcripts in TB, before combining these to form a discriminating multi-gene signature. Simplifying these to single gene biomarkers has the added benefit of facilitating their integration into panels of blood RNA biomarkers for multinomial classification^39^ that may overcome the specificity limitations of the current binomial approach, for example by combining with the best performing single gene biomarkers of viral infections^40^.

An important strength of our analysis is that we have adopted ICE-TB terminology for subclinical TB, to facilitate comparisons across studies and biomarker domains. We included 6530 samples including 283 samples from subclinical TB cases, making it the largest analysis of RNA biomarkers for subclinical TB to date. Our data processing pipeline ensured batch correction within studies and integrated RNAseq and qPCR data into a pooled dataset. We also performed the first decision curve analysis, to our knowledge, to quantify the clinical utility of RNA biomarkers for subclinical TB and compare to existing tests. We additionally stratified our decision curve analysis by TB burden, which allowed more granular assessment of clinical utility by setting. We also performed a number of sensitivity analyses, including a two-stage IPD-MA, to ensure our primary findings were robust.

A limitation of our study is that, whilst we included cohorts from high and low burden settings, there were few contributing countries (South Africa, UK, Brazil, The Gambia, and Ethiopia) with no representation from Asia, although large proportions of the UK studies were individuals of South Asian ethnicity. There were variations in case definitions and baseline screening between studies which may have resulted in misclassification, however this is reflective of real-world variations in clinical practice according to resource availability. Furthermore, with the exception of CORTIS-01/HR, evaluation of TB disease during follow-up was symptom-triggered so additional cases of subclinical TB may have been missed. There were low numbers of subclinical cases over a 12-month interval in some studies, with both Leicester Contacts and CORTIS-HR reporting 5 cases each, however this reflects the reality that TB is a relatively rare outcome in longitudinal cohort studies. We also acknowledge that there is no gold standard for the ‘subclinical, non-infectious’ state and high-resolution investigations for macroscopic pathology, such as PET-CT^41^, were not performed. We therefore assumed that participants who developed TB within 12 months would have had macroscopic pathology at baseline, had high-resolution investigation been performed. However ongoing *Mtb* exposure during follow-up, particularly in high transmission settings, may mean that disease cases within the primary 12-month interval may be attributable to new infection and may have led to underestimation of sensitivity for subclinical TB. Since subclinical TB may regress or undulate without treatment^42^, it is also possible that we underestimated specificity for ‘subclinical, non-infectious’ TB that did not progress to clinical disease within 12 months. Future studies will be required to further evaluate the accuracy of candidate biomarkers for the ‘subclinical, non-infectious’ state, once a scalable and widely accepted reference standard is established.

In summary, we have demonstrated that several single-gene transcripts perform equivalently to multi-gene signatures to detect subclinical TB, which may simplify assays and encourage commercial competition. RNA biomarker performance is consistent across settings and exceeds performance of IGRA in high burden settings, but falls short of WHO benchmarks. A combination strategy with IGRA shows promise to enable more targeted preventative treatment in low incidence settings.

## Footnotes

## Acknowledgements

We thank the study teams and participants from all of the primary studies included in this analysis (ACS, CORTIS-01, CORTIS-HR, GC6-74, Leicester Contacts, London Contacts and REPORT-Brazil).

## Author contributions

JGB, MN, and RKG conceived the study. JGB and RKG wrote the analysis plan with input from all authors. JGB performed the analyses and wrote the first draft of the manuscript, supported by RKG and MN. All authors contributed to the methods and interpretation and approved the final submitted manuscript.

## Data availability

All data analysed in this study are publicly available through the original contributing studies.

