## Supplementary for "Single-gene transcripts for subclinical TB: an individual participant data meta-analysis"

Supplementary Appendix

### Supplementary Methods

#### *Systematic search*

We searched PubMed with no language or date restrictions using the following search strategy:

*(tuberculosis[Title/Abstract]) AND (subclinical[Title/Abstract] OR incipient[Title/Abstract] OR progression[Title/Abstract]) AND (rna[Title/Abstract] OR transcript*[Title/Abstract])*

We included original studies where participants of any age underwent whole blood RNA sampling with at least 12 months of follow-up for development of clinical TB. We included studies containing participants living with HIV and those receiving preventative therapy. Studies that used genome-wide (RNAseq or microarray) or targeted transcriptional profiling (qPCR or NanoString quantification) were included.

#### *RNA data processing pipeline*

The qPCR data processing pipeline for the REPORT-Brazil and CORTIS-01/HR datasets are shown in supplementary figure 1. Individual samples with more than 20% failed primer probes were excluded. Where <20% of probes failed, the sample was included for all available transcripts, as per the original contributing studies.

For the REPORT-Brazil dataset, we normalised genes by subtracting the individual raw gene cycle thresholds from the mean cycle threshold of three reference genes (TMBIM6, USF2, ACTR3). We identified batch effects between the qPCR chips (supplementary figure 3E) and so we performed batch-correction using quantile regression, including the TB outcome in the model, using the *batchtma* package in R[2]. There was a subsequent improvement in batch effects (supplementary figure 3F).

In the CORTIS-01/HR studies, two parallel assays were run, labelled .x and .y. The .x assay contained four reference genes (TMBIM6, CDC42, USF2, ACTR3). The .y assay did not contain reference genes however paired probes were present in both assays (GBP1.Hs00977005_m1, GBP2.Hs00894846_g1, GBP5.Hs00369472_m1, SERPING1.Hs00934329_m1). We first normalised .x genes by subtracting individual raw gene cycle thresholds from the mean cycle threshold of the reference genes, before we batch-corrected the normalised .x genes (as described for REPORT-Brazil), including .x reference genes in order to avoid introducing batch effects into the .y assay. We then normalised the .y genes by subtracting the raw gene cycle thresholds from the mean of the batch-corrected .x reference genes. We calculated an additional corrective factor using the difference between the normalised paired probes measured in both assays before adding this corrective factor to the normalised .y genes. There was an increase in correlation of .x and .y primers after this normalisation step (supplementary figure 2). Finally we performed batch-correction of the .y genes. Density plots demonstrated an improvement in batch effects (supplementary figures 3A-D).

Where there were multiple primers per gene, we explored correlation between primers. Since primers for each gene were highly correlated (supplementary figure 6), we selected the primer with the least missingness for each gene within each study (supplementary figure 5). Expression of FCGR1A and its duplicated pseudogene FCGR1B[31] were used interchangeably because measurements in the available studies do not reliably distinguish between these transcripts. Since FCGR1B was available across all studies, this was used to represent FCGR1A/B.

We included eight parsimonious signatures that we included in our previous analysis, selected pragmatically based on availability of previous validation data, number of constituent genes and availability of target sequences[3]. The eight included signatures consisted of 2-11 constituent genes; Francisco2, Roe3, Sweeney3, Maetzdorf4, Suliman4, Thompson5, Penn-Nicholson6, and Darboe11. For signatures that required machine learning methods, we used the difference of means of the upregulated genes minus the means of the downregulated genes as a simplified approach to calculation to improve reproducibility, since previous analyses have demonstrated similar performance against original methods[30]. The signatures calculated using genes in the CORTIS .y assay did not require normalisation as these signatures contained both upregulated and downregulated genes, and so the non-normalised gene values were used to calculate signatures. It was not possible to calculate Darboe11 for participants from the REPORT-Brazil dataset as four of the constituent genes were absent from the qPCR panel.

We included 80 single-genes that were present in the RNAseq dataset and at least one qPCR study (CORTIS-01/HR or REPORT-Brazil [supplementary figure 5A]).

We standardised signatures and single-gene transcripts within each RNAseq and qPCR dataset by converting to z-scores, which allowed us to pool qPCR and RNAseq datasets. We used non-progressors to calculate means and standard deviations in each contributing dataset. Prior to calculation of means and standard deviations, we winsorized non-progressors to remove extreme outliers (below the 1st or above the 99th percentile) using the *Winsorize* function in R[4]. As CORTIS-HR included only people living with HIV, we used the non-progressors from CORTIS-01 to calculate z-scores for CORTIS-HR. Density plots of gene expression stratified by dataset before and after z-score transformation suggest that this method was effective in aligning the data across studies (supplementary figures 4A & 4B).

***References***

1 Gupta RK, Turner CT, Venturini C, *et al.* Concise whole blood transcriptional signatures for incipient tuberculosis: a systematic review and patient-level pooled meta-analysis. *The Lancet Respiratory Medicine* 2020; **8**: 395–406.

2 Stopsack, K, Gerke, T. batchtma: Batch Effect Adjustments. https://stopsack.github.io/batchtma.

3 Mendelsohn SC, Mbandi SK, Fiore-Gartland A, *et al.* Prospective multicentre head-to-head validation of host blood transcriptomic biomarkers for pulmonary tuberculosis by real-time PCR. *Commun Med* 2022; **2**: 1–13.

4 Signorell A. Winsorize function - RDocumentation. https://www.rdocumentation.org/packages/DescTools/versions/0.99.54/topics/Winsorize (accessed June 24, 2024).

5 Viechtbauer W. Conducting Meta-Analyses in R with the metafor Package. *Journal of Statistical Software* 2010; **36**: 1–48.

**Supplementary figures & tables: methods**

***Supplementary figure 1: Flowchart of data processing pipeline***

Flowchart demonstrating the data processing pipeline for the qPCR studies. In CORTIS, parallel assays (.x and .y) were performed and so the processing pipeline differed.

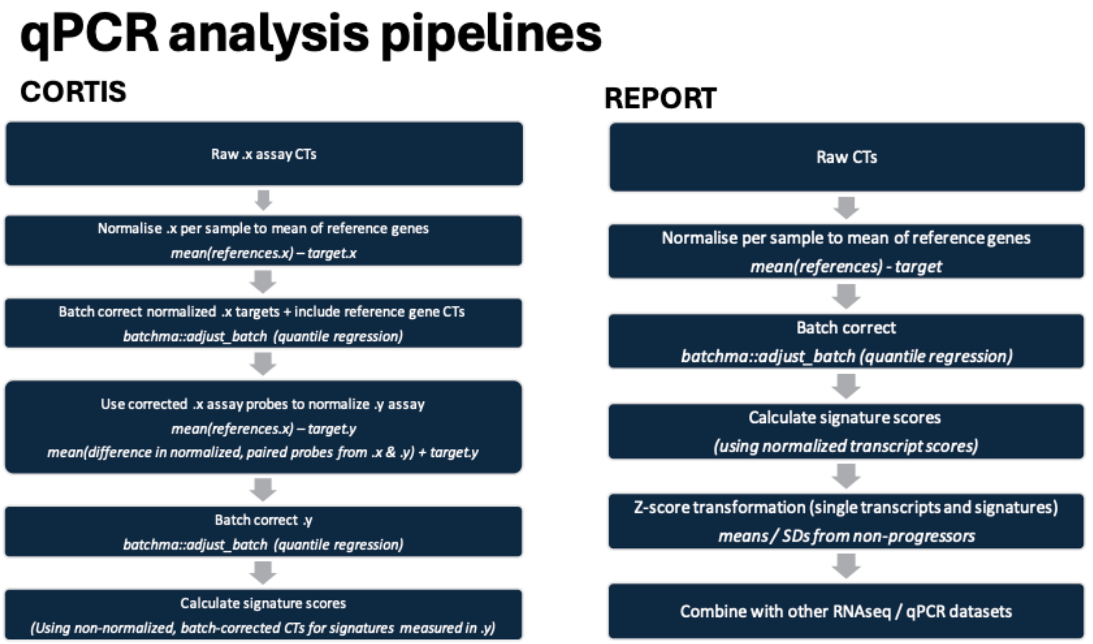

***Supplementary figure 2:  Normalisation of CORTIS .y primers***

Correlation heatmap of .x and .y primers in CORTIS, shown pre-normalisation (left) and post-normalisation (right) of .y primers. Since reference genes were not included in the .y assays, normalisation of .y primers was performed by first normalising to batch-corrected .x reference genes and then using a corrective factor calculated as the mean difference between paired .x and .y primers. Correlation between .x and .y primers improved markedly following normalization.

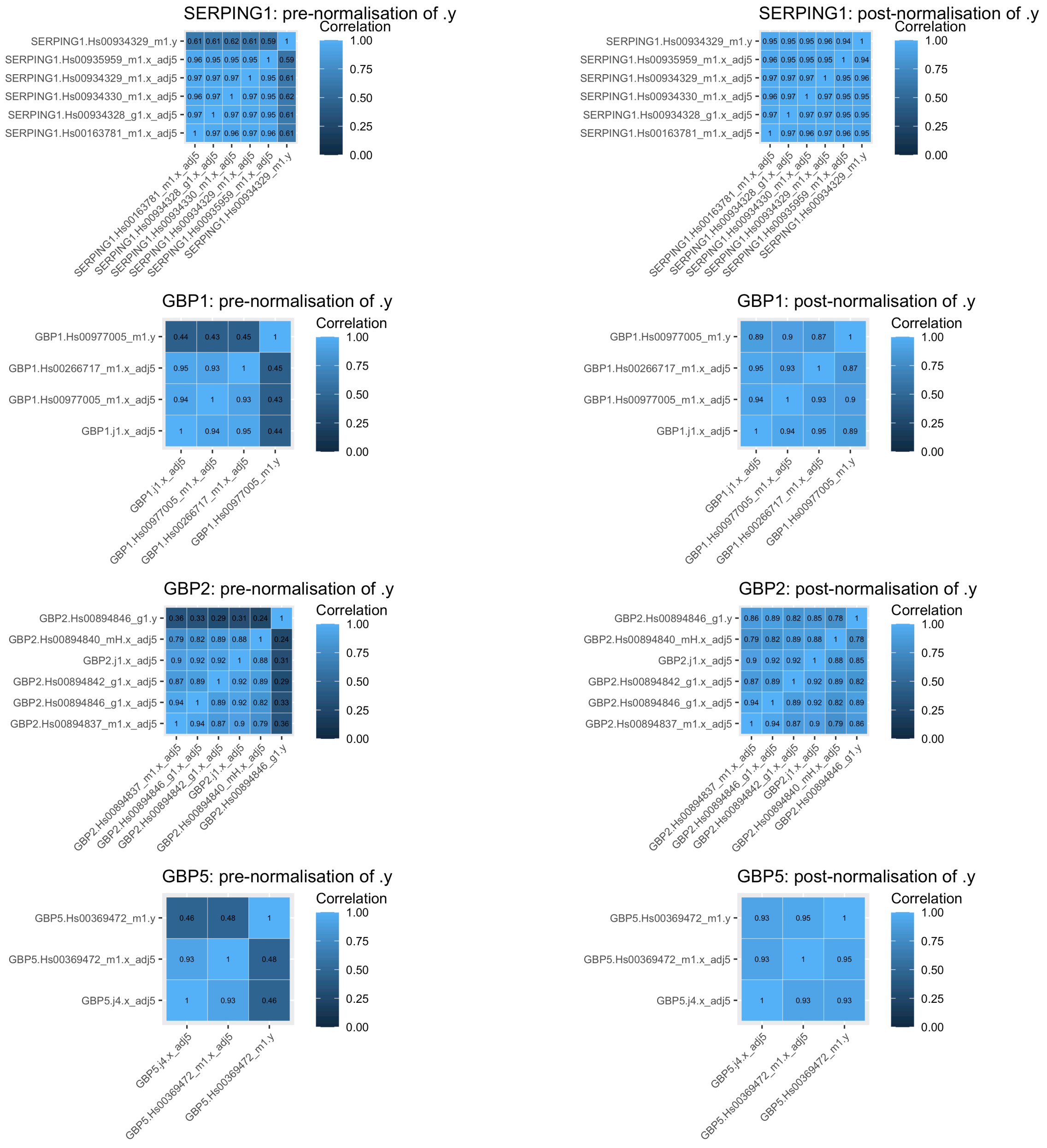

***Supplementary figure 3: Batch correction***

Density plots of gene expression, separated by qPCR batch, are shown before and after batch correction for nine randomly selected CORTIS .x, CORTIS .y and REPORT-Brazil primers. Batch correction was performed using quantile regression, including the TB outcome in the model (*batchtma::adjust_batch*).

***Supplementary figure 3A: Density plots of gene expression in CORTIS .x primers, stratified by PCR batch, before batch correction***

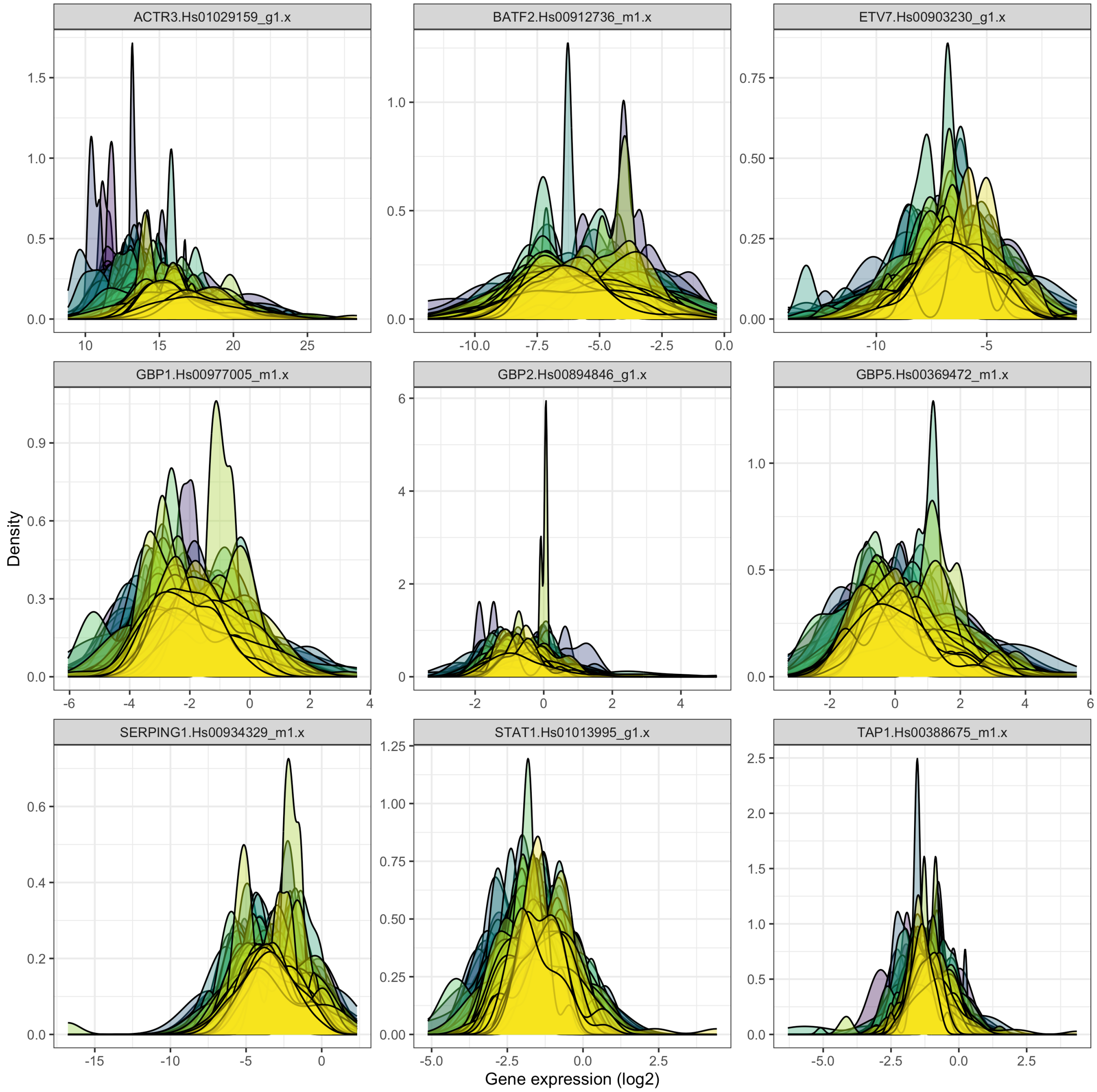

***Supplementary figure 3B: Density plots of gene expression in CORTIS .x primers, stratified by PCR batch, after batch correction***

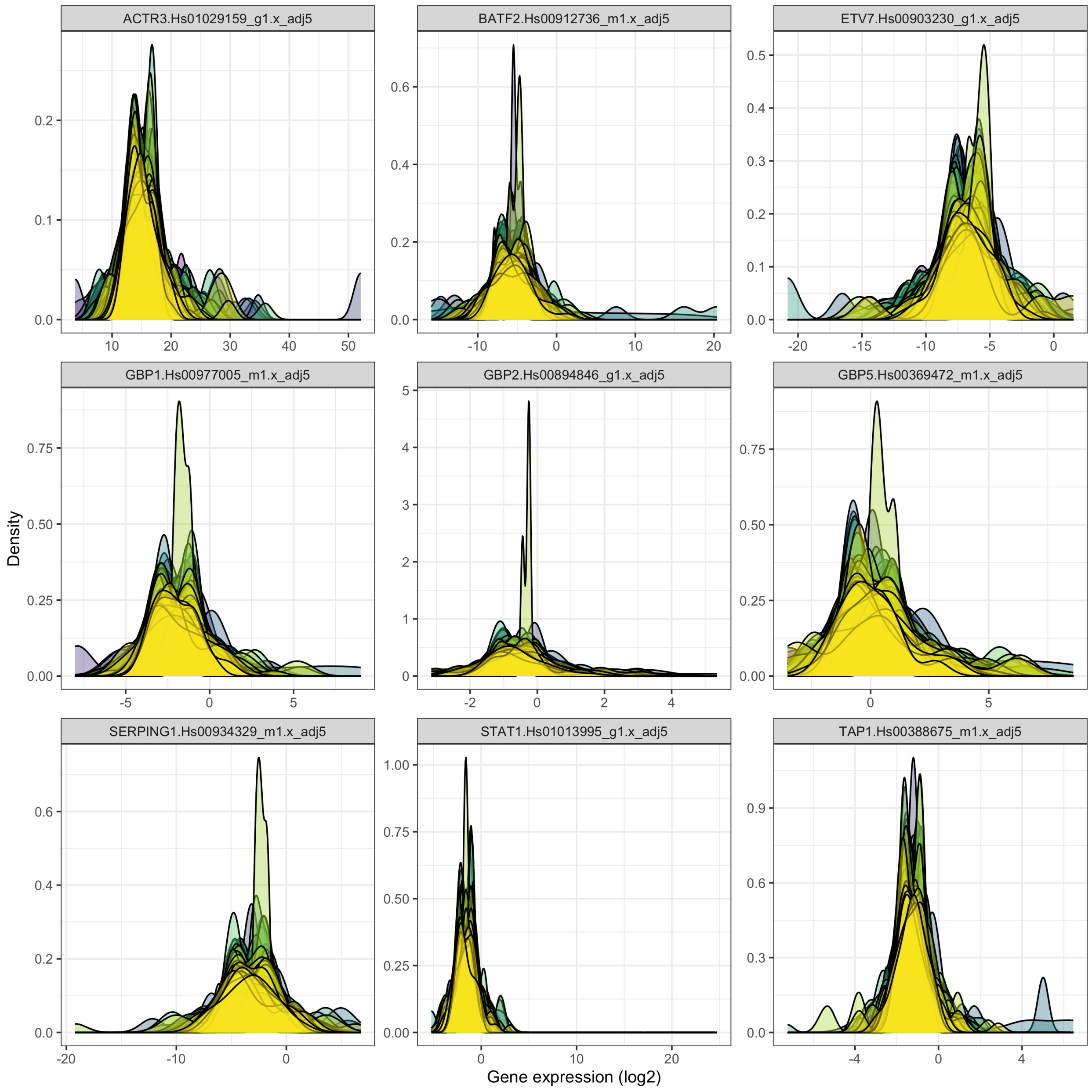

***Supplementary figure 3C: Density plots of gene expression in CORTIS .y primers, stratified by PCR batch, before batch correction***

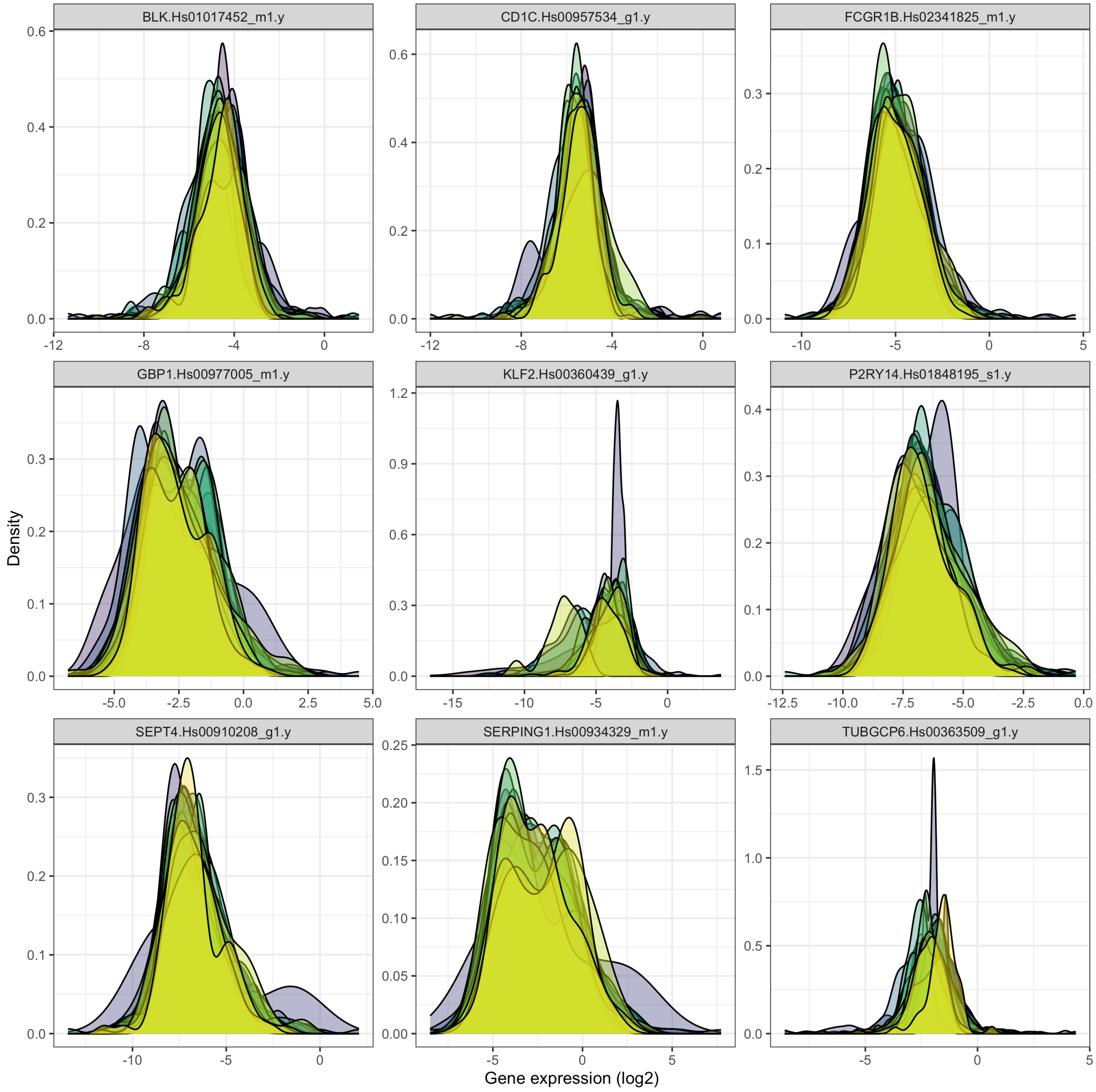

***Supplementary figure 3D: Density plots of gene expression in CORTIS .y primers, stratified by PCR batch, after batch correction***

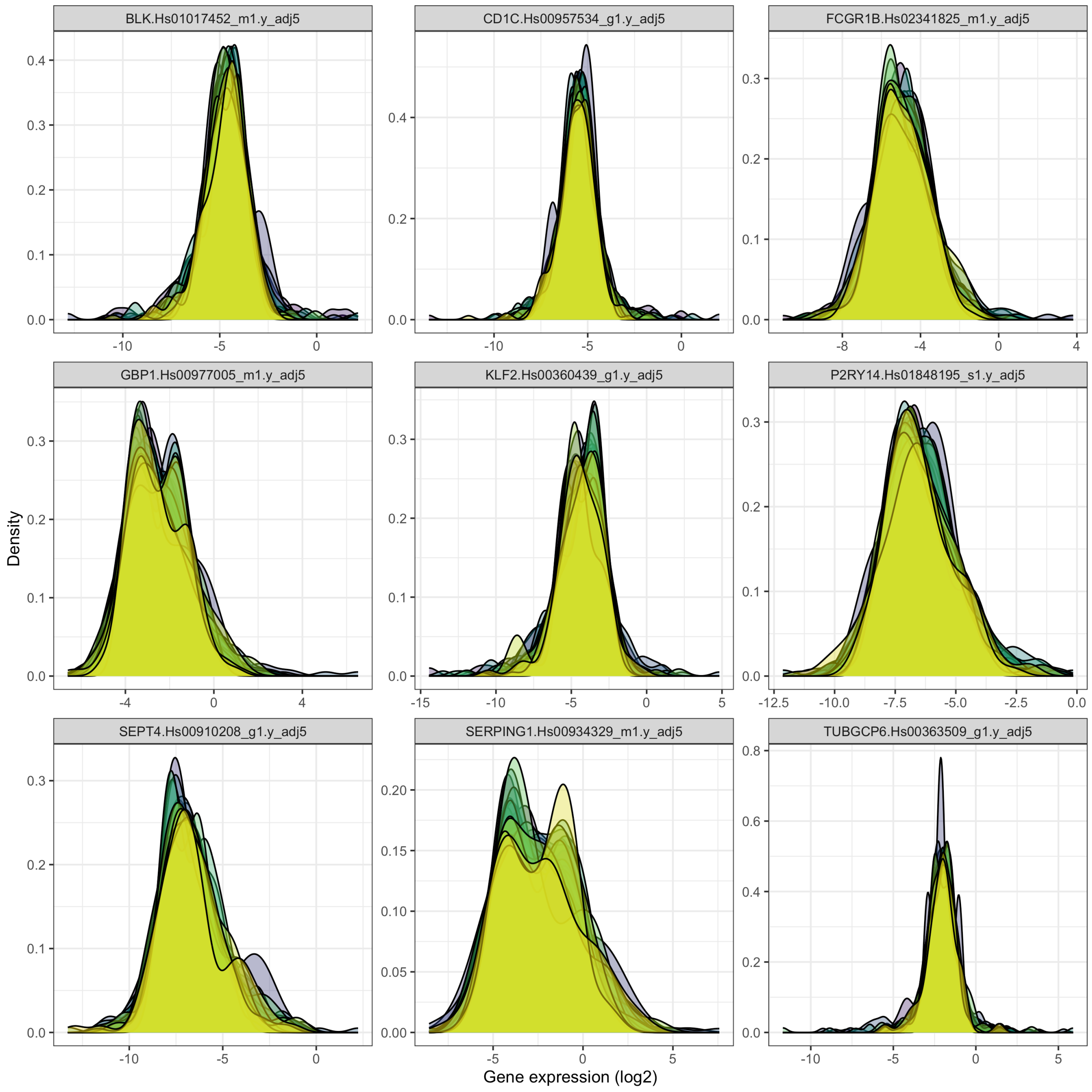

***Supplementary figure 3E: Density plots of gene expression in REPORT-Brazil, stratified by PCR batch, before batch correction***

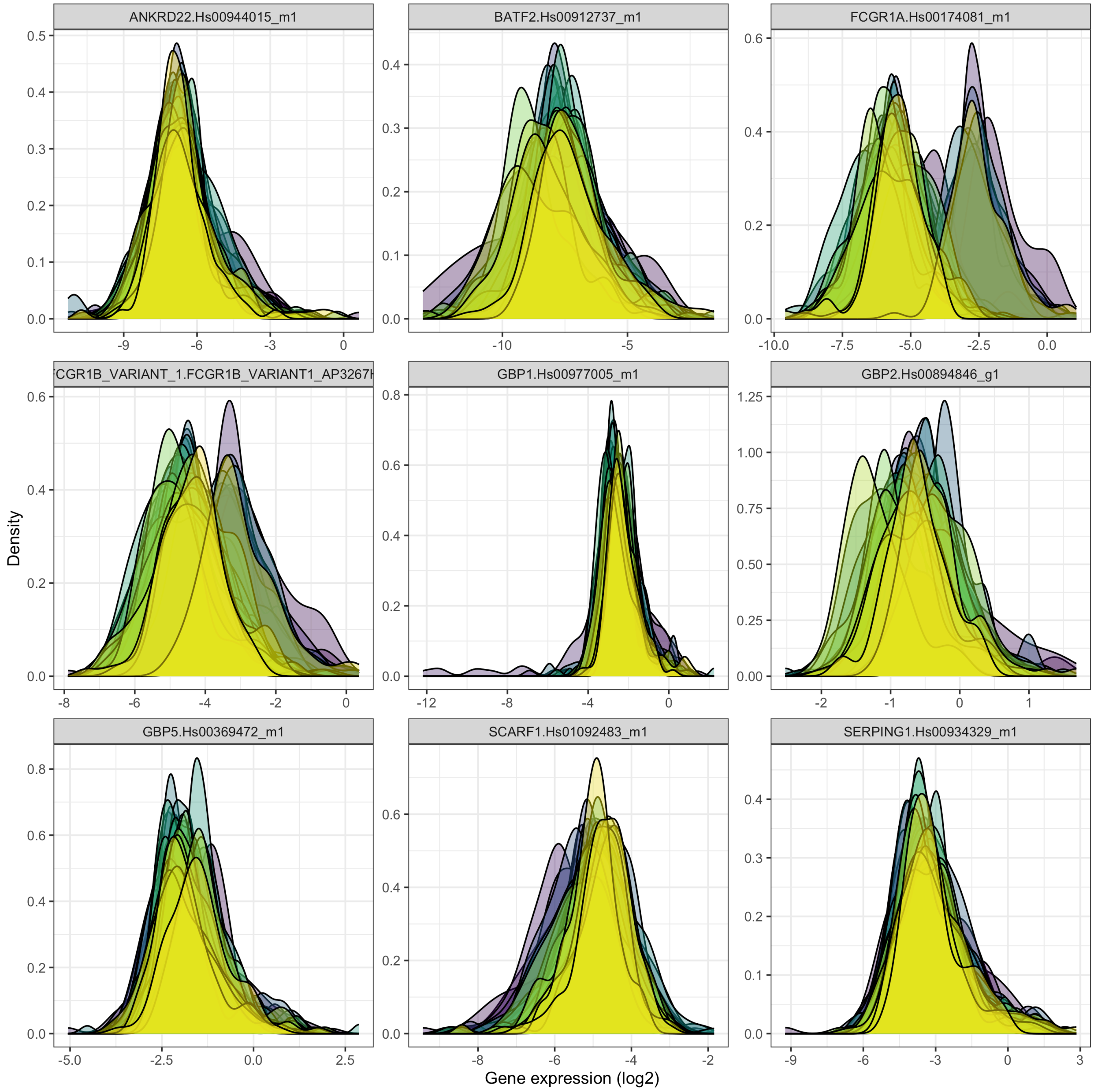

***Supplementary figure 3F: Density plots of gene expression in REPORT-Brazil, stratified by PCR batch, after batch correction***

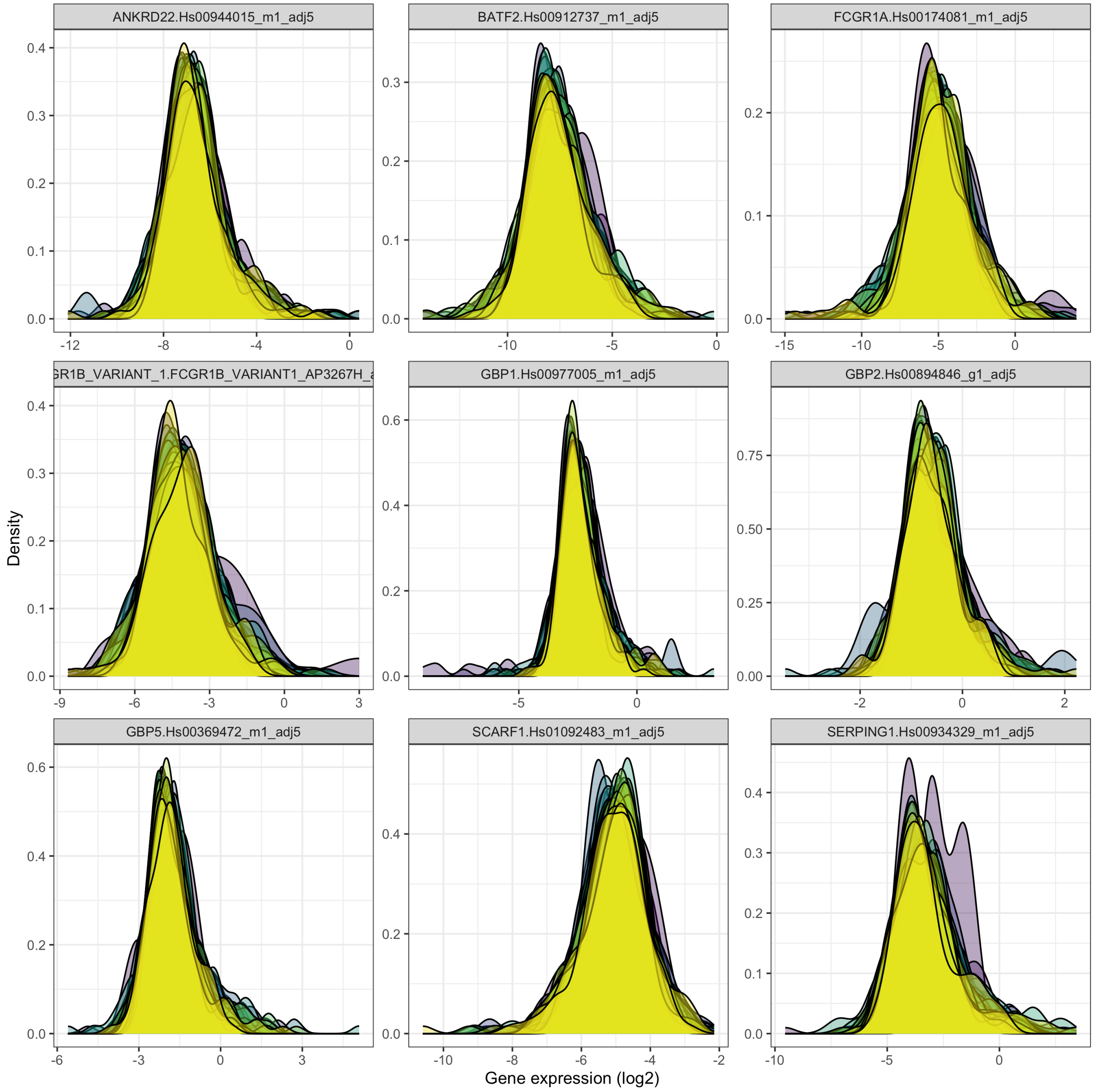

***Supplementary figure 4: Z-score transformation***

Density plots of gene expression, separated by cohort, are shown before and after z-score transformation. Z-score transformation was performed separately for the RNAseq studies, CORTIS and REPORT-Brazil using a winsorized cohort of healthy non-progressors to calculate means and standard deviation, using HIV-negative, non-progressors from CORTIS-01 for both CORTIS-01 and CORTIS-HR.

***Supplementary figure 4A: Density plots of expression of single-gene transcripts present in all studies, stratified by study, before z-score transformation***

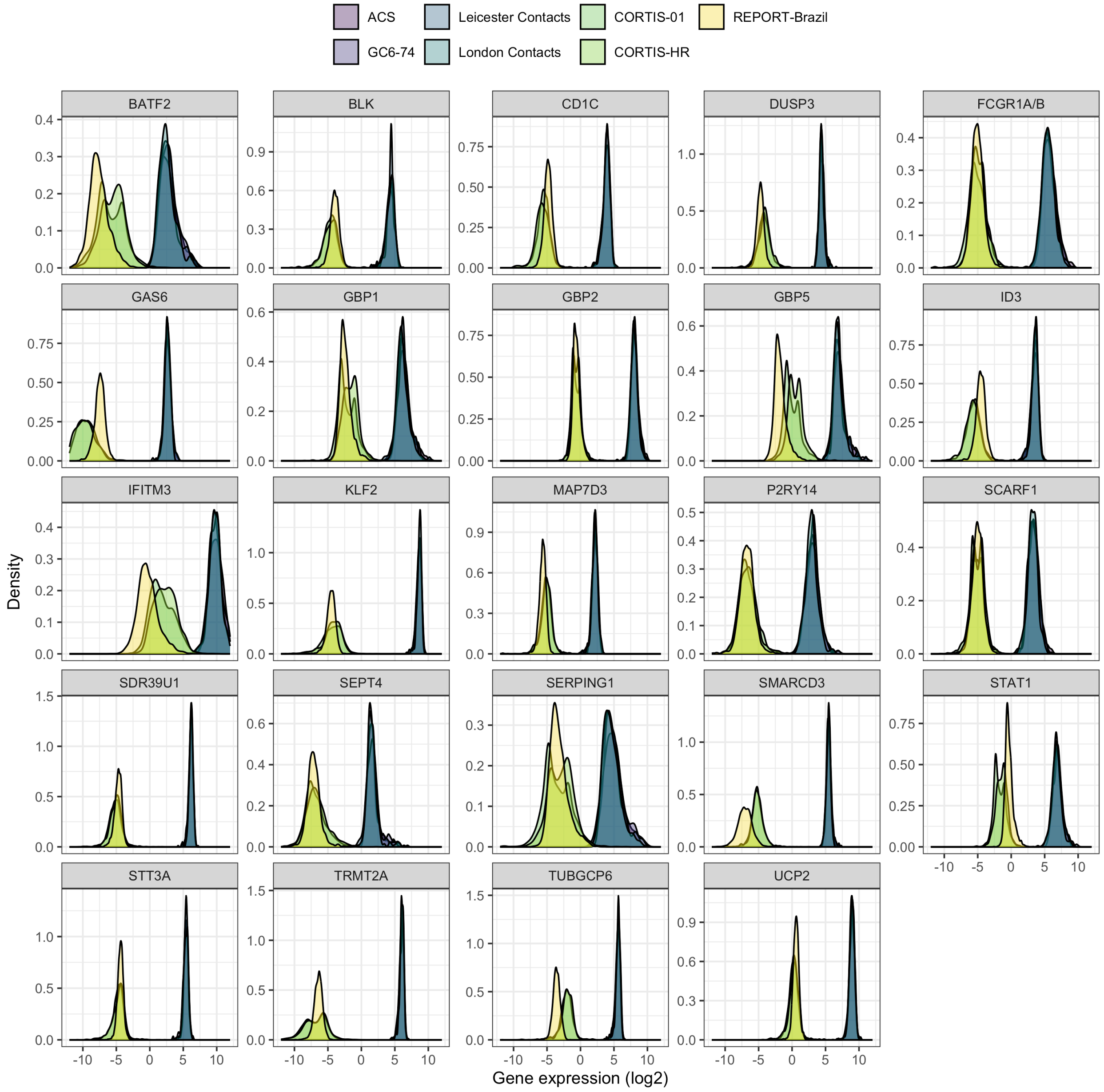

***Supplementary figure 4B: Density plots of expression of single-gene transcripts present in all studies, stratified by study, after z-score transformation***

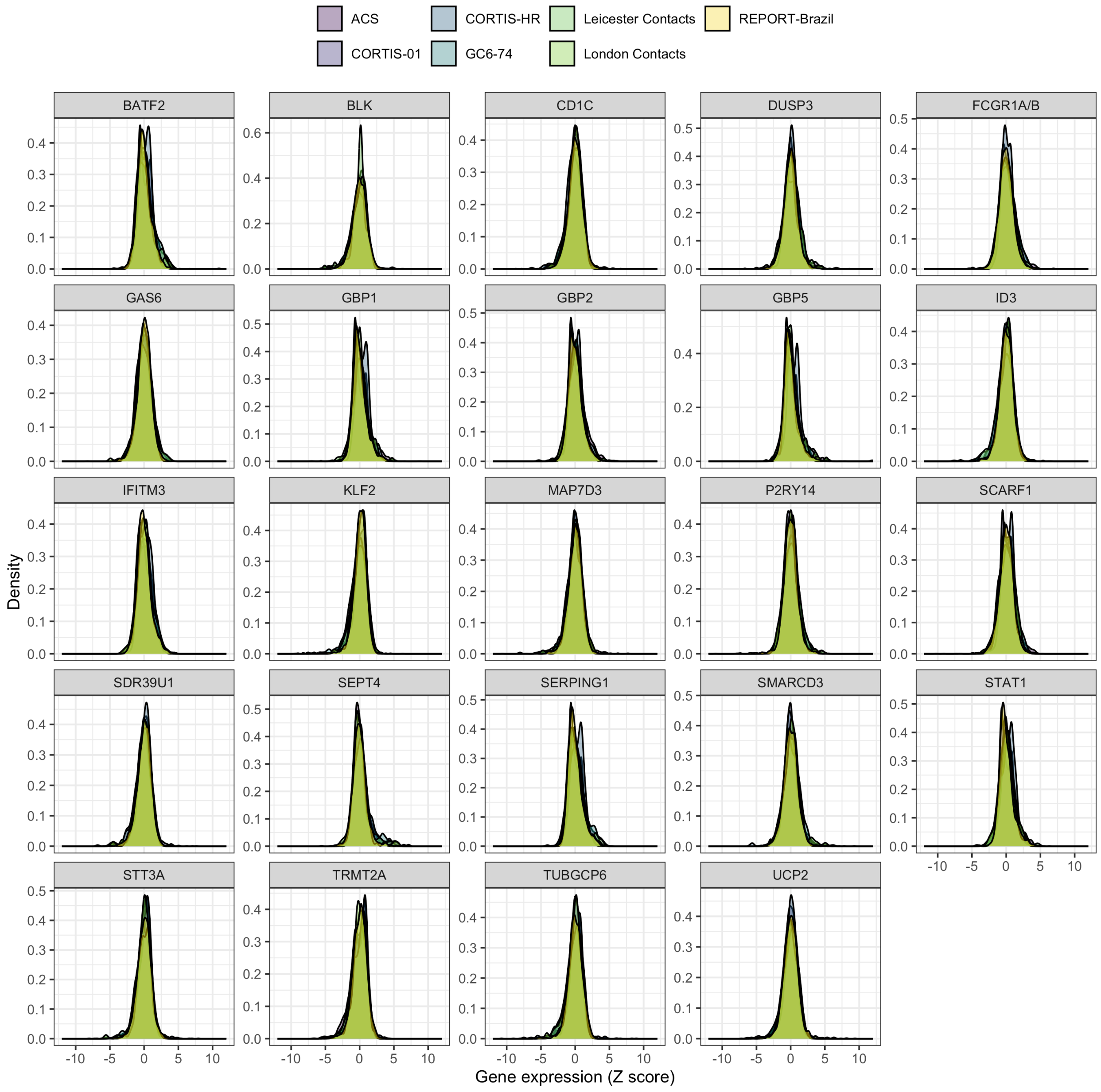

***Supplementary figure 5: Gene missingness***

***Supplementary figure 5A: Missingness plot of genes, stratified by study***

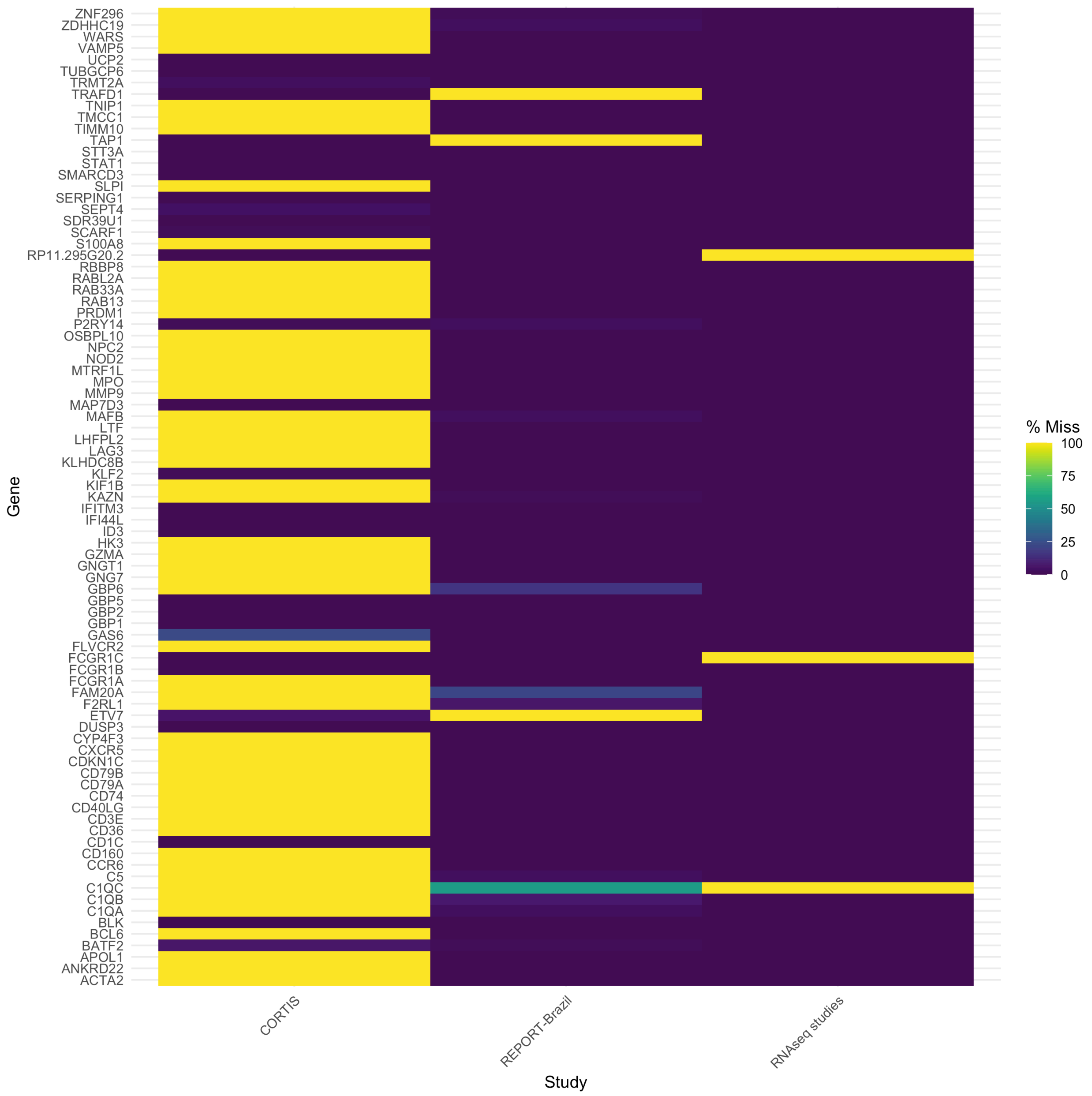

***Supplementary figure 5B: Missingness plot of primers for individual genes from CORTIS-01, stratified by TB status***

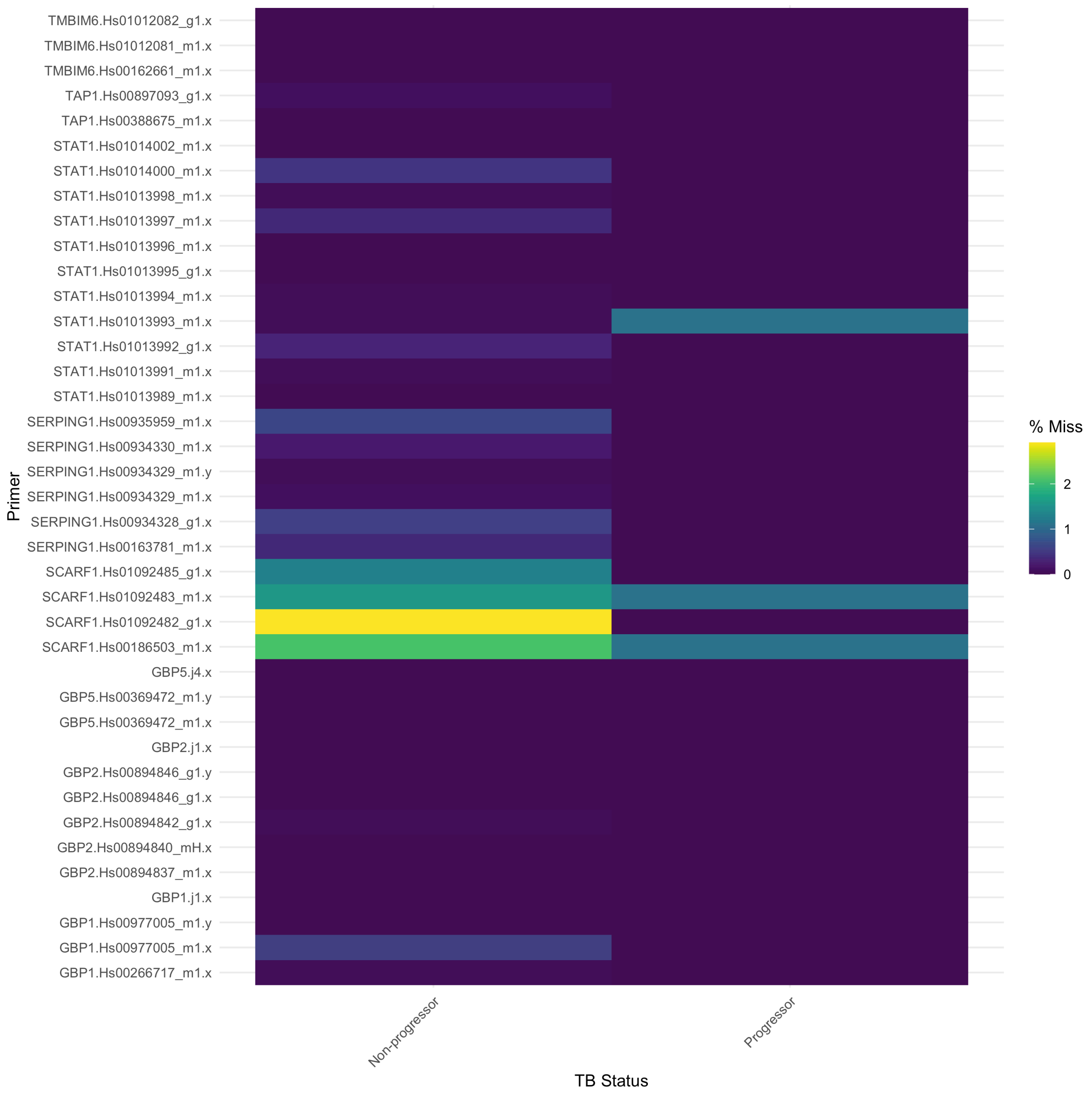

***Supplementary figure 5C: Missingness plot of primers for individual genes from CORTIS-HR, stratified by TB status***

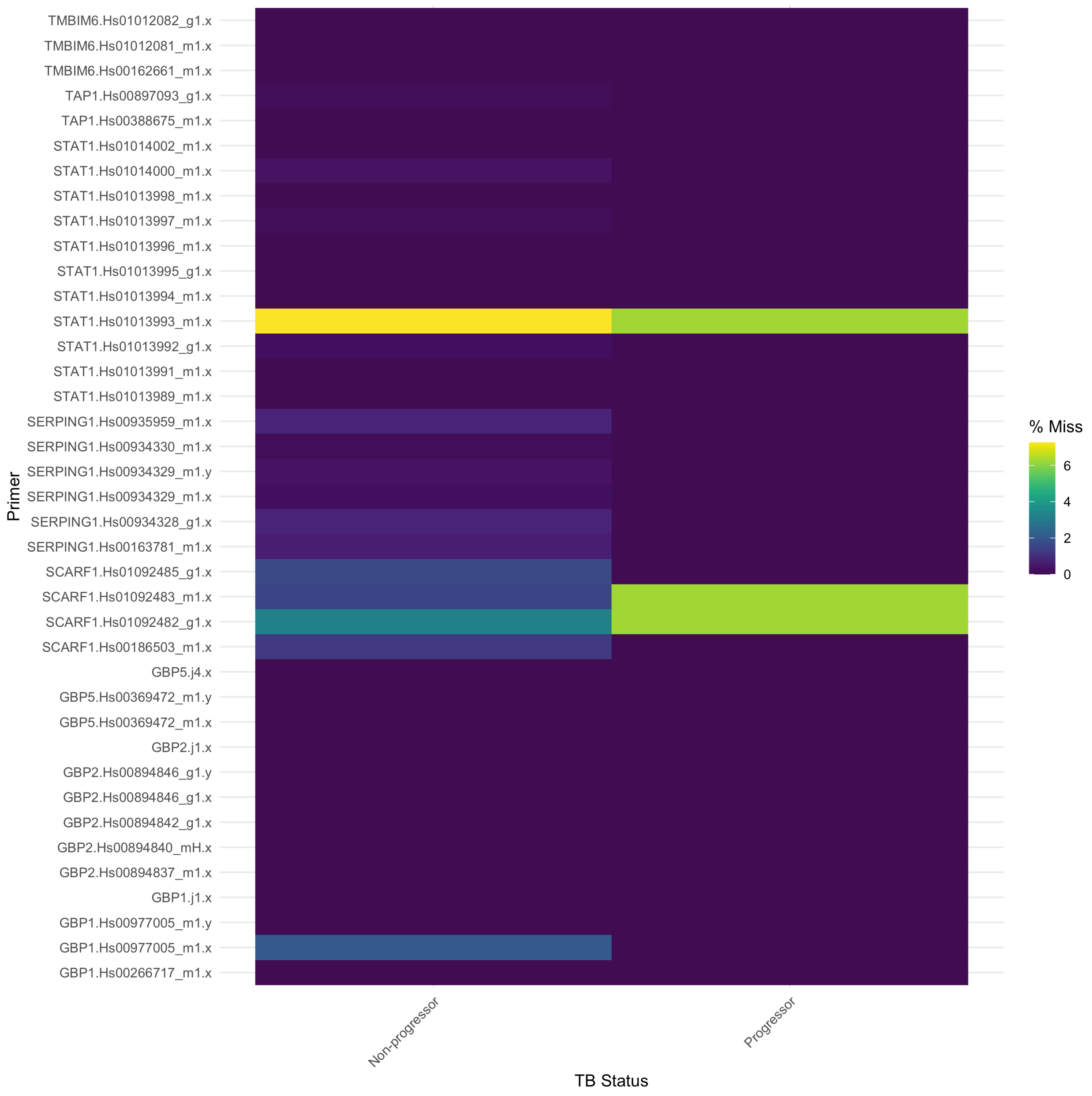

***Supplementary figure 5D: Missingness plot of primers for individual genes from REPORT-Brazil, stratified by TB status***

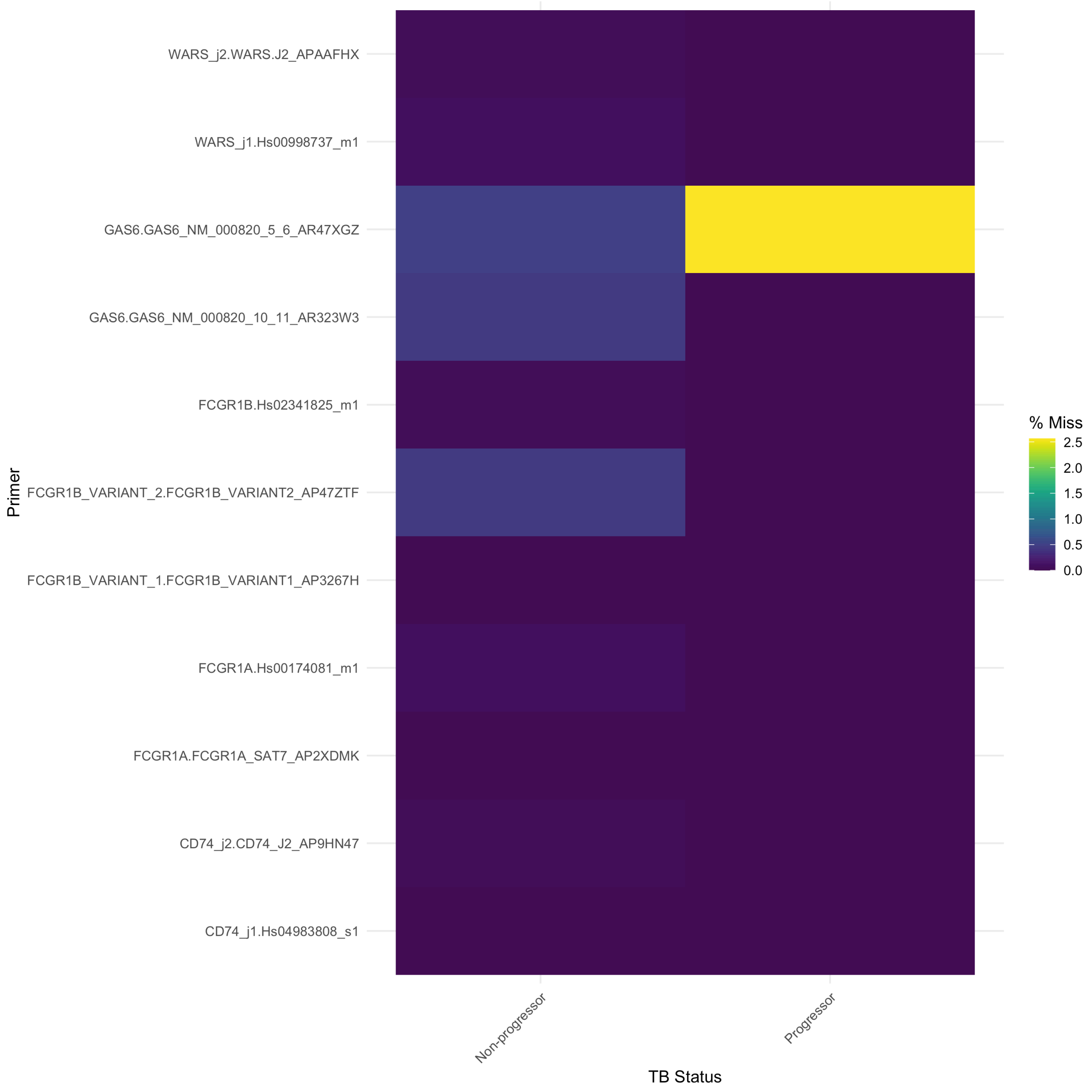

***Supplementary figure 6: Correlation of multiple primers for individual genes***

***Supplementary figure 6A: Correlation heatmaps of multiple primers for individual genes in CORTIS-01/HR***

Correlation heatmap of genes in CORTIS-01/HR where multiple primers per gene were available, with primers clustered by gene. Scale is shown from 0 to 1. As primers for individual genes were highly correlated, the primer with the least missingness was used to represent each gene.

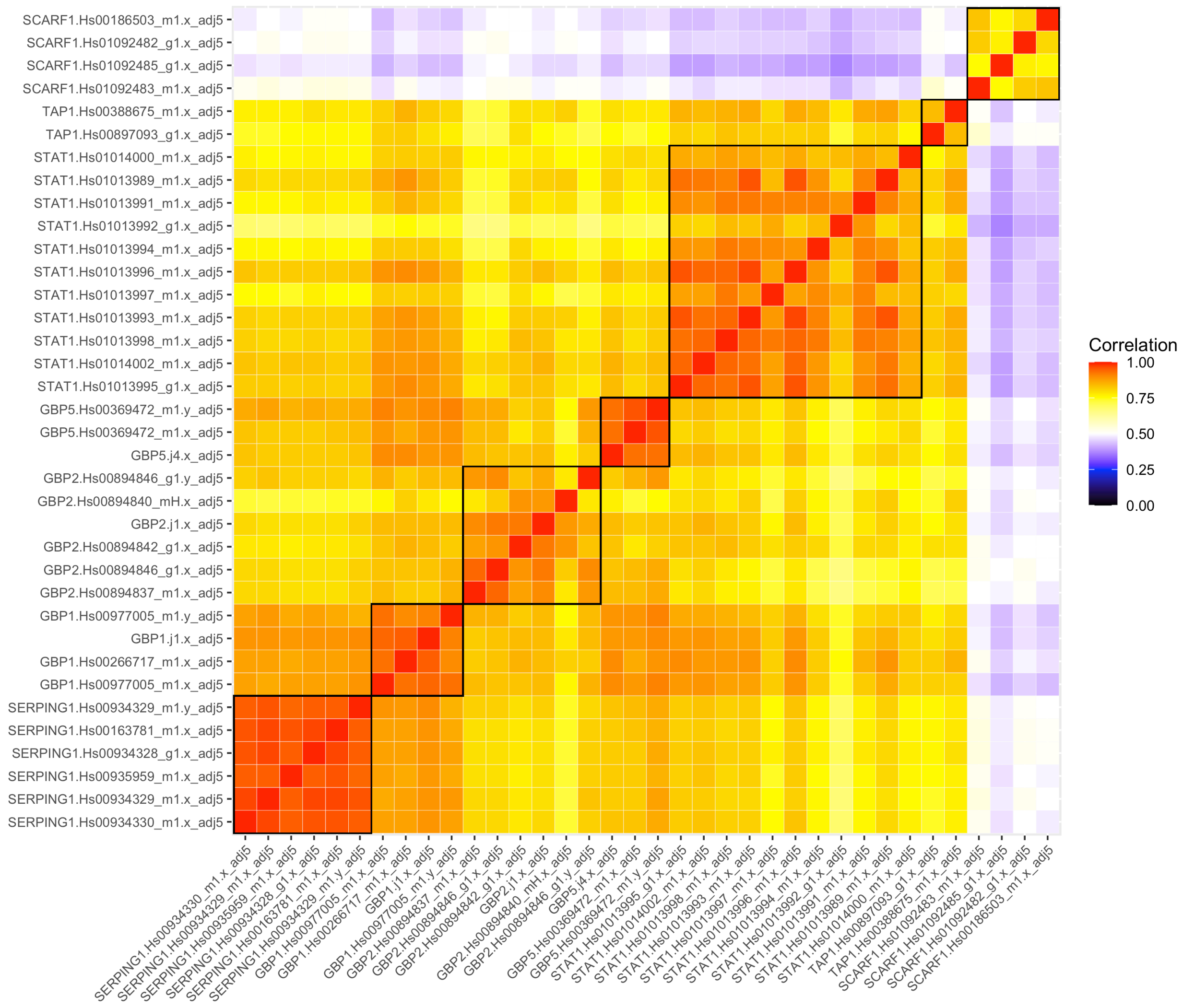

***Supplementary figure 6B: Correlation heatmaps of multiple primers for individual genes in REPORT-Brazil***

Correlation heatmap of genes in REPORT-Brazil where multiple primers per gene were available, with primers clustered by gene. Scale is shown from -1 to 1. As primers for individual genes were highly correlated, the primer with the least missingness was used to represent each gene.

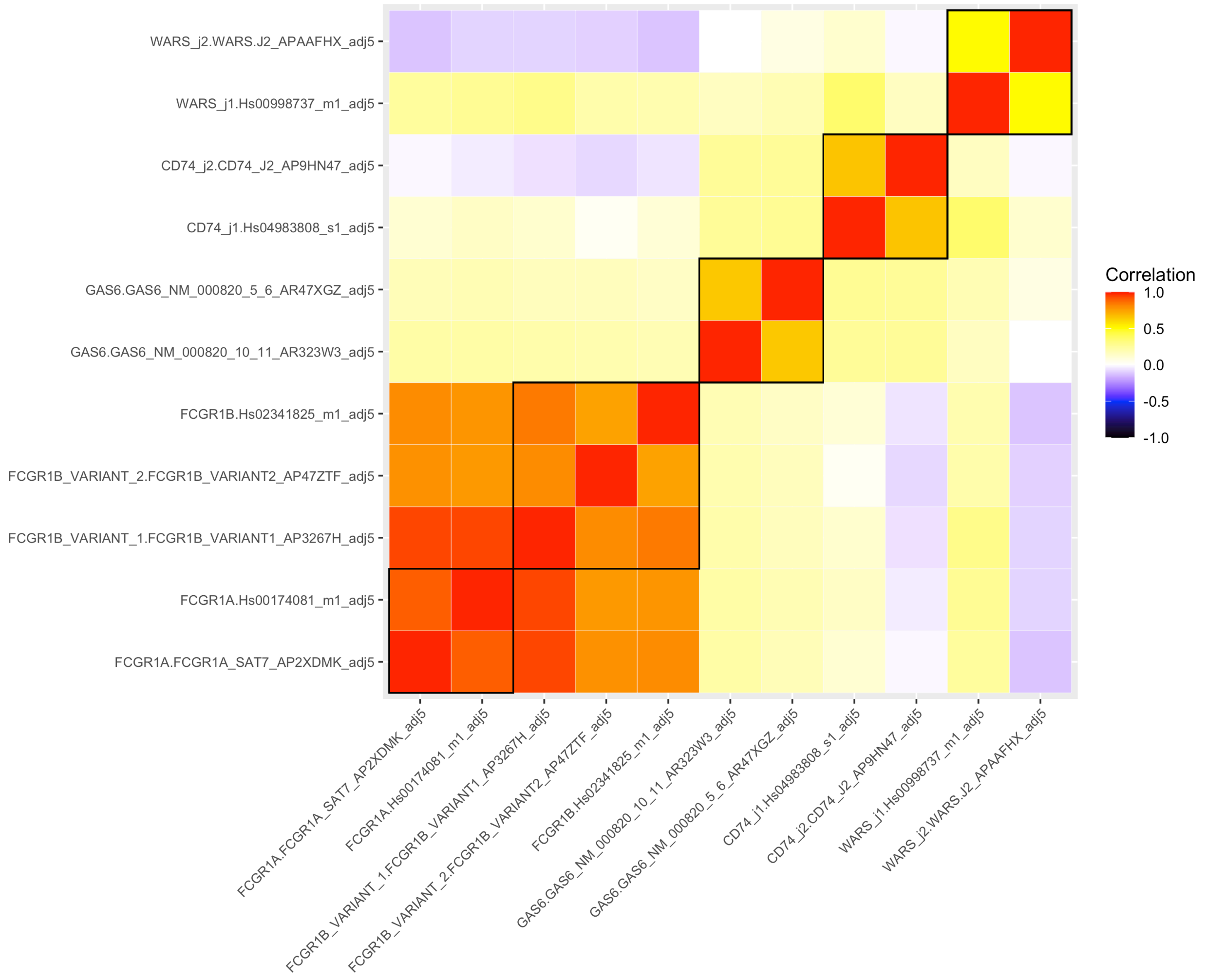

***Supplementary figure 7: Intra- and inter-individual variance in serial samples (RNA and IGRA)***

A plot of intraclass correlation (ICC) of transcripts and IGRA for serial samples from one individual. ICC is the proportion of total variance due to inter-individual variation. Here ICC was calculated using two methods; mixed effects and variance components. ICC ratios of >0.5, as seen for IGRA, means that interindividual variation contributes more to total variance than intraindividual variation and so serial samples from one individual correlate and were not considered as independent. ICC ratios of ≤0.5, as seen for most transcripts, means that interindividual variation contributes to total variance equally to or less than intraindividual variation and so serial samples from one individual were considered as independent.

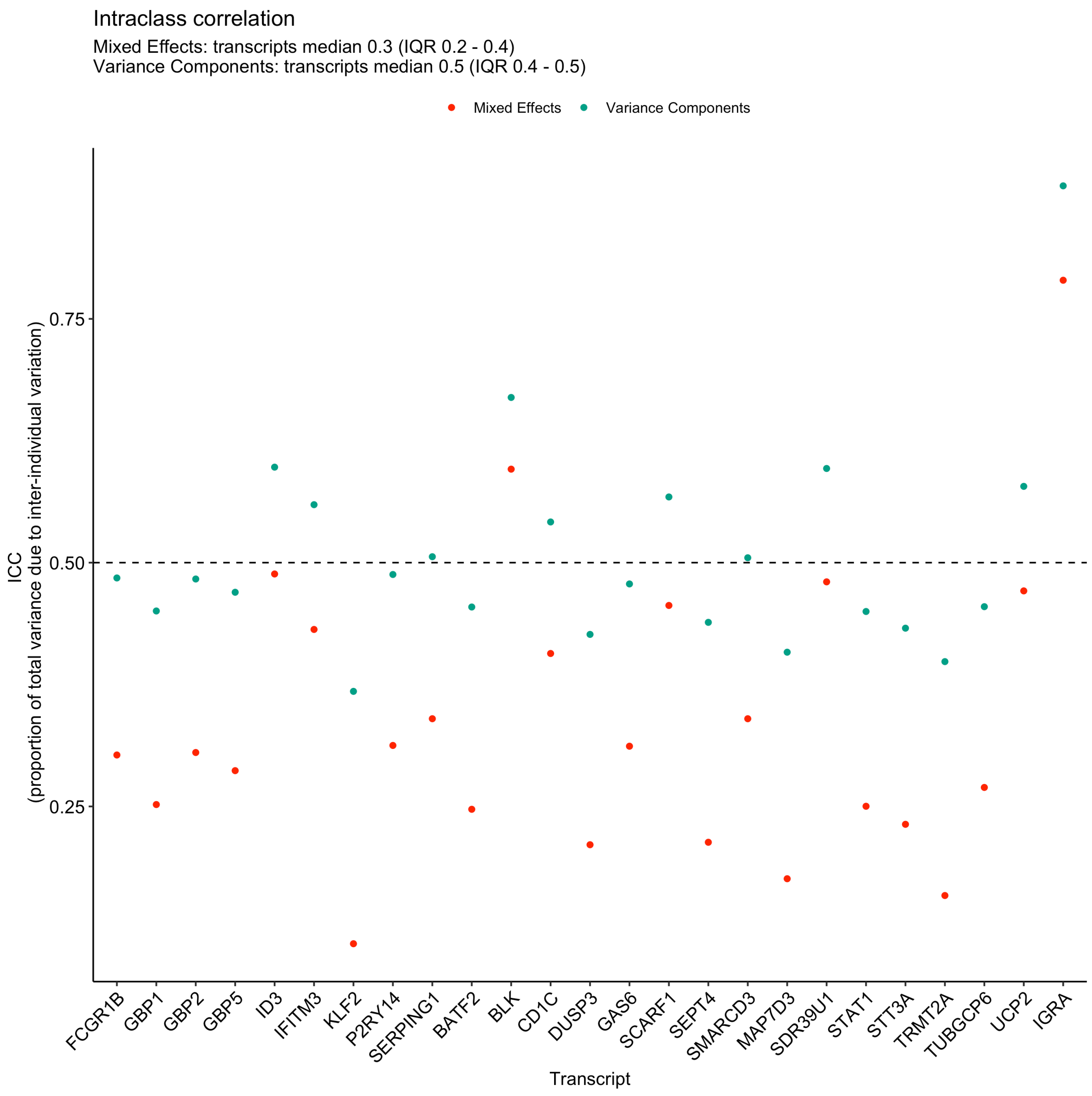

### Supplementary figures & tables: results

#### *Supplementary figure 8: Flowchart of included samples from contributing datasets in meta-analysis*

A flowchart of number of RNA samples and individuals from each contributing dataset that were included in the meta-analysis. Number of excluded samples and reasons for exclusion are also shown. *Indicates >1 sample collected from the same participant within a 6-month interval.

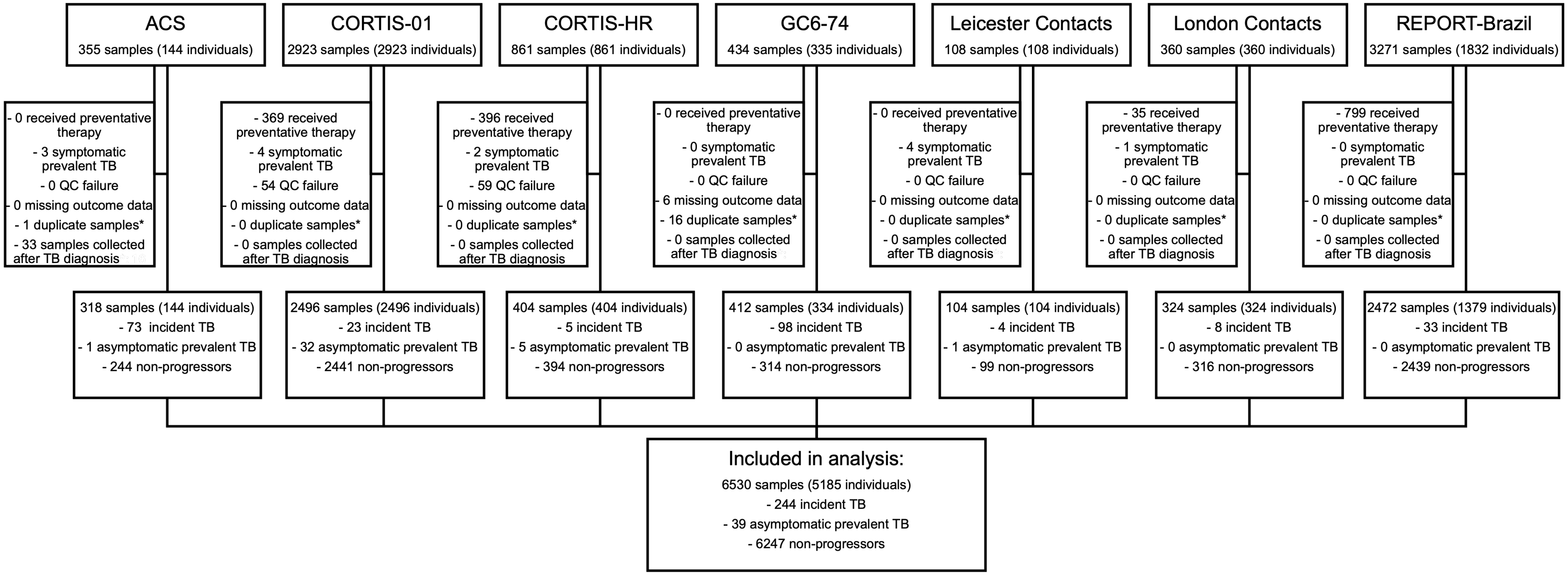

***Supplementary table 1: Table of participant characteristics by study***

A table of characteristics of samples included in the primary analysis (excluding TPT recipients) overall and by study.

| **Characteristic** | **Overall, N = 6,544[1]** | **ACS, N = 321[1]** | **GC6-74, N = 412[1]** | **Leicester Contacts, N = 108[1]** | **London Contacts, N = 325[1]** | **CORTIS-01, N = 2,500[1]** | **CORTIS-HR, N = 406[1]** | **REPORT-Brazil, N = 2,472[1]** |
|---|---|---|---|---|---|---|---|---|
| **Age** | 26 (21-35) | 16 (15-17) | 23 (18-35) | 35 (25-46) | 34 (26-47) | 26 (22-33) | 35 (29-41) | NA (NA-NA) |
| **Sex** | | | | | | | | |
| **Female** | 2,110 (58%) | 228 (71%) | 239 (58%) | 0 (NA%) | 0 (NA%) | 1,347 (54%) | 296 (73%) | 0 (NA%) |
| **Male** | 1,529 (42%) | 93 (29%) | 173 (42%) | 0 (NA%) | 0 (NA%) | 1,153 (46%) | 110 (27%) | 0 (NA%) |
| **Clinical TB (symptomatic prevalent cases)** | 14 (0.2%) | 3 (1.4%) | 0 (0%) | 4 (3.9%) | 1 (0.3%) | 4 (0.2%) | 2 (0.5%) | 0 (0%) |
| **Subclinical TB, infectious (asymptomatic prevalent cases)** | 39 (0.6%) | 1 (0.5%) | 0 (0%) | 1 (1.0%) | 0 (0%) | 32 (1.3%) | 5 (1.3%) | 0 (0%) |
| **Subclinical TB, non-infectious (incident cases)** | 244 (3.8%) | 73 (25%) | 98 (24%) | 4 (3.9%) | 8 (2.5%) | 23 (0.9%) | 5 (1.3%) | 33 (1.3%) |
| **Subclinical TB, total** | 283 (4.4%) | 74 (26%) | 98 (24%) | 5 (4.8%) | 8 (2.5%) | 55 (2.2%) | 10 (2.5%) | 33 (1.3%) |
| **Months 0-6** | 115 | 15 | 47 | 5 | 2 | 35 | 5 | 6 |
| **Months 6-12** | 75 | 28 | 19 | 0 | 5 | 11 | 0 | 12 |
| **Months 12-15** | 44 | 13 | 9 | 0 | 0 | 9 | 5 | 8 |
| **Months 15+** | 49 | 18 | 23 | 0 | 1 | 0 | 0 | 7 |

[1]Median (25%-75%); n (%); n

***Supplementary figure 9: Correlation heatmaps of best performing single-gene transcripts***

Spearman rank correlation heatmap of single-gene transcripts with equivalent performance to the best performing multi-gene signature (Roe3).

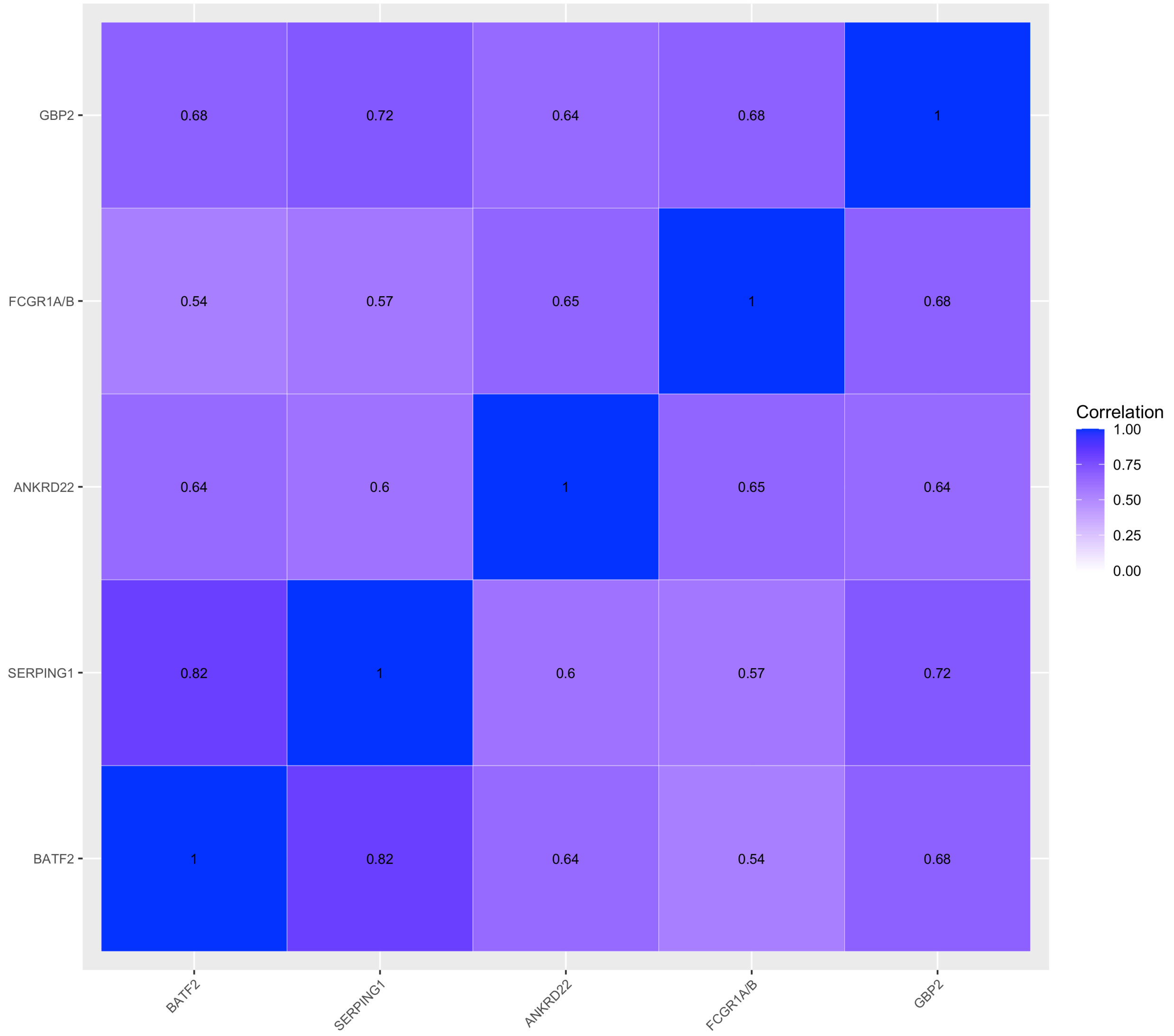

***Supplementary table 2: Performance metrics table of all single-gene transcripts to discriminate subclinical TB from non-progressors at various intervals (0-12 months, 0-3 months, 0-6 months, 0-15 months, 6-12 months, and 12-15 months)***

Area Under the Receiver Operating Curve (AUROC) estimates and 95% CIs of all single-gene transcripts and the best performing multi-gene signature (Roe3) to discriminate subclinical TB cases from non-progressors at various time intervals from sampling to disease are shown in this table. Equivalence to Roe3, the best multi-gene signature, was defined as an adjusted p value >0.05 in pairwise Delong test for 0-12 months.

| | **0-12 months (primary analysis)** | | | | **0-3 months** | | **0-6 months** | | **0-15 months** | | **6-12 months** | | **12-15 months** | |
|---|---|---|---|---|---|---|---|---|---|---|---|---|---|---|
| **Signature** | **AUROC** | **Cases** | **Controls** | **p** | **AUROC** | **Cases** | **AUROC** | **Cases** | **AUROC** | **Cases** | **AUROC** | **Cases** | **AUROC** | **Cases** |
| **Roe3** | 0.77 (0.73 - 0.81) | 189 | 4,982 | | 0.81 (0.76 - 0.86) | 73 | 0.78 (0.73 - 0.83) | 115 | 0.74 (0.71 - 0.78) | 232 | 0.76 (0.69 - 0.82) | 74 | 0.61 (0.52 - 0.7) | 43 |
| **BATF2** | 0.77 (0.73 - 0.81) | 189 | 4,982 | 0.715 | 0.81 (0.76 - 0.86) | 73 | 0.78 (0.73 - 0.82) | 115 | 0.74 (0.7 - 0.77) | 232 | 0.75 (0.69 - 0.82) | 74 | 0.6 (0.51 - 0.69) | 43 |
| **FCGR1A/B** | 0.77 (0.73 - 0.81) | 189 | 4,981 | 0.867 | 0.8 (0.75 - 0.86) | 73 | 0.77 (0.72 - 0.81) | 115 | 0.75 (0.71 - 0.78) | 232 | 0.77 (0.71 - 0.83) | 74 | 0.66 (0.57 - 0.75) | 43 |
| **ANKRD22** | 0.77 (0.72 - 0.81) | 138 | 3,243 | 0.881 | 0.85 (0.78 - 0.92) | 34 | 0.77 (0.71 - 0.83) | 75 | 0.74 (0.7 - 0.79) | 168 | 0.77 (0.7 - 0.83) | 63 | 0.63 (0.52 - 0.74) | 30 |
| **GBP2** | 0.75 (0.71 - 0.79) | 189 | 4,982 | 0.064 | 0.78 (0.72 - 0.83) | 73 | 0.75 (0.7 - 0.8) | 115 | 0.73 (0.69 - 0.76) | 232 | 0.75 (0.69 - 0.81) | 74 | 0.62 (0.53 - 0.71) | 43 |
| **GBP5** | 0.75 (0.71 - 0.79) | 189 | 4,982 | 0.012 | 0.79 (0.73 - 0.85) | 73 | 0.75 (0.7 - 0.8) | 115 | 0.72 (0.68 - 0.76) | 232 | 0.74 (0.68 - 0.8) | 74 | 0.59 (0.5 - 0.69) | 43 |
| **SERPING1** | 0.75 (0.71 - 0.79) | 189 | 4,979 | 0.069 | 0.77 (0.72 - 0.83) | 73 | 0.76 (0.71 - 0.81) | 115 | 0.74 (0.7 - 0.77) | 232 | 0.74 (0.68 - 0.8) | 74 | 0.65 (0.57 - 0.74) | 43 |
| **GBP1** | 0.74 (0.7 - 0.78) | 189 | 4,968 | 0.006 | 0.77 (0.71 - 0.83) | 73 | 0.74 (0.69 - 0.79) | 115 | 0.72 (0.68 - 0.76) | 232 | 0.74 (0.68 - 0.81) | 74 | 0.61 (0.52 - 0.69) | 43 |
| **ETV7** | 0.73 (0.69 - 0.77) | 172 | 2,531 | 0.032 | 0.73 (0.67 - 0.79) | 68 | 0.73 (0.68 - 0.78) | 109 | 0.72 (0.68 - 0.76) | 207 | 0.72 (0.65 - 0.79) | 63 | 0.66 (0.55 - 0.76) | 35 |
| **APOL1** | 0.73 (0.68 - 0.78) | 138 | 3,243 | 0.005 | 0.78 (0.69 - 0.88) | 34 | 0.75 (0.68 - 0.81) | 75 | 0.71 (0.66 - 0.75) | 168 | 0.71 (0.63 - 0.78) | 63 | 0.59 (0.49 - 0.7) | 30 |
| **TAP1** | 0.71 (0.67 - 0.76) | 172 | 2,588 | 0.002 | 0.73 (0.66 - 0.79) | 68 | 0.71 (0.66 - 0.77) | 109 | 0.7 (0.66 - 0.74) | 207 | 0.71 (0.65 - 0.78) | 63 | 0.63 (0.52 - 0.73) | 35 |
| **SCARF1** | 0.71 (0.67 - 0.75) | 189 | 4,982 | 0.000 | 0.75 (0.69 - 0.81) | 73 | 0.73 (0.67 - 0.78) | 115 | 0.69 (0.65 - 0.73) | 232 | 0.68 (0.62 - 0.75) | 74 | 0.61 (0.51 - 0.7) | 43 |
| **STAT1** | 0.71 (0.67 - 0.75) | 189 | 4,982 | 0.000 | 0.73 (0.67 - 0.79) | 73 | 0.71 (0.67 - 0.76) | 115 | 0.69 (0.66 - 0.73) | 232 | 0.71 (0.64 - 0.78) | 74 | 0.59 (0.5 - 0.68) | 43 |

| | 0-12 months (primary analysis) | | | | 0-3 months | | 0-6 months | | 0-15 months | | 6-12 months | | 12-15 months | |
|---|---|---|---|---|---|---|---|---|---|---|---|---|---|---|
| Signature | AUROC | Cases | Controls | p | AUROC | Cases | AUROC | Cases | AUROC | Cases | AUROC | Cases | AUROC | Cases |
| **C1QB** | 0.71 (0.66 - 0.76) | 137 | 3,087 | 0.012 | 0.8 (0.71 - 0.89) | 33 | 0.75 (0.69 - 0.81) | 74 | 0.69 (0.64 - 0.74) | 166 | 0.67 (0.6 - 0.75) | 63 | 0.57 (0.46 - 0.68) | 29 |
| **SEPT4** | 0.71 (0.66 - 0.75) | 186 | 4,927 | 0.000 | 0.73 (0.67 - 0.8) | 72 | 0.72 (0.66 - 0.77) | 114 | 0.68 (0.64 - 0.72) | 229 | 0.69 (0.62 - 0.76) | 72 | 0.58 (0.49 - 0.68) | 43 |
| **C1QA** | 0.7 (0.65 - 0.75) | 138 | 3,192 | 0.003 | 0.78 (0.69 - 0.87) | 34 | 0.73 (0.67 - 0.79) | 75 | 0.67 (0.63 - 0.72) | 168 | 0.66 (0.58 - 0.74) | 63 | 0.56 (0.45 - 0.66) | 30 |
| **LHFPL2** | 0.7 (0.65 - 0.75) | 138 | 3,229 | 0.000 | 0.75 (0.64 - 0.85) | 34 | 0.69 (0.62 - 0.77) | 75 | 0.69 (0.64 - 0.73) | 168 | 0.7 (0.63 - 0.77) | 63 | 0.62 (0.52 - 0.72) | 30 |
| **NPC2** | 0.7 (0.65 - 0.75) | 138 | 3,239 | 0.005 | 0.81 (0.73 - 0.88) | 34 | 0.73 (0.67 - 0.79) | 75 | 0.68 (0.64 - 0.73) | 168 | 0.67 (0.59 - 0.74) | 63 | 0.59 (0.48 - 0.7) | 30 |
| **DUSP3** | 0.69 (0.65 - 0.74) | 189 | 4,982 | 0.000 | 0.76 (0.7 - 0.82) | 73 | 0.71 (0.66 - 0.76) | 115 | 0.67 (0.64 - 0.71) | 232 | 0.67 (0.6 - 0.74) | 74 | 0.59 (0.5 - 0.68) | 43 |
| **CXCR5** | 0.69 (0.64 - 0.74) | 138 | 3,244 | 0.010 | 0.78 (0.69 - 0.86) | 34 | 0.71 (0.65 - 0.78) | 75 | 0.67 (0.63 - 0.72) | 168 | 0.66 (0.59 - 0.73) | 63 | 0.6 (0.48 - 0.72) | 30 |
| **IFITM3** | 0.68 (0.64 - 0.72) | 189 | 4,982 | 0.000 | 0.7 (0.64 - 0.76) | 73 | 0.69 (0.64 - 0.74) | 115 | 0.67 (0.63 - 0.7) | 232 | 0.68 (0.61 - 0.74) | 74 | 0.57 (0.48 - 0.67) | 43 |
| **P2RY14** | 0.68 (0.64 - 0.72) | 189 | 4,878 | 0.000 | 0.71 (0.64 - 0.78) | 73 | 0.67 (0.62 - 0.73) | 115 | 0.66 (0.62 - 0.7) | 232 | 0.68 (0.62 - 0.75) | 74 | 0.58 (0.49 - 0.67) | 43 |
| **FLVCR2** | 0.68 (0.63 - 0.73) | 138 | 3,227 | 0.000 | 0.78 (0.69 - 0.87) | 34 | 0.69 (0.62 - 0.76) | 75 | 0.66 (0.61 - 0.7) | 168 | 0.67 (0.59 - 0.75) | 63 | 0.47 (0.35 - 0.58) | 30 |
| **VAMP5** | 0.68 (0.63 - 0.73) | 138 | 3,244 | 0.002 | 0.79 (0.69 - 0.89) | 34 | 0.69 (0.62 - 0.77) | 75 | 0.66 (0.62 - 0.71) | 168 | 0.67 (0.6 - 0.74) | 63 | 0.58 (0.47 - 0.69) | 30 |
| **OSBPL10** | 0.68 (0.63 - 0.72) | 138 | 3,230 | 0.004 | 0.69 (0.59 - 0.79) | 34 | 0.69 (0.62 - 0.75) | 75 | 0.67 (0.63 - 0.72) | 168 | 0.66 (0.58 - 0.73) | 63 | 0.65 (0.55 - 0.75) | 30 |
| **FAM20A** | 0.67 (0.62 - 0.73) | 135 | 2,755 | 0.000 | 0.79 (0.71 - 0.87) | 33 | 0.68 (0.61 - 0.75) | 74 | 0.65 (0.6 - 0.7) | 164 | 0.67 (0.59 - 0.74) | 61 | 0.46 (0.35 - 0.58) | 29 |
| **CD79A** | 0.67 (0.62 - 0.72) | 138 | 3,244 | 0.002 | 0.69 (0.59 - 0.78) | 34 | 0.69 (0.62 - 0.75) | 75 | 0.66 (0.61 - 0.71) | 168 | 0.64 (0.57 - 0.72) | 63 | 0.63 (0.52 - 0.73) | 30 |
| **GBP6** | 0.67 (0.62 - 0.72) | 135 | 2,874 | 0.000 | 0.77 (0.68 - 0.85) | 33 | 0.68 (0.61 - 0.75) | 74 | 0.64 (0.59 - 0.68) | 165 | 0.65 (0.58 - 0.72) | 61 | 0.5 (0.42 - 0.59) | 30 |
| **TIMM10** | 0.67 (0.62 - 0.72) | 138 | 3,244 | 0.000 | 0.78 (0.7 - 0.85) | 34 | 0.69 (0.62 - 0.76) | 75 | 0.65 (0.6 - 0.69) | 168 | 0.64 (0.57 - 0.71) | 63 | 0.56 (0.44 - 0.67) | 30 |
| **SMARCD3** | 0.67 (0.62 - 0.71) | 189 | 4,980 | 0.000 | 0.73 (0.67 - 0.8) | 73 | 0.7 (0.64 - 0.75) | 115 | 0.66 (0.61 - 0.7) | 232 | 0.62 (0.55 - 0.69) | 74 | 0.61 (0.5 - 0.71) | 43 |

| | 0-12 months (primary analysis) | | | | 0-3 months | | 0-6 months | | 0-15 months | | 6-12 months | | 12-15 months | |
|---|---|---|---|---|---|---|---|---|---|---|---|---|---|---|
| **Signature** | **AUROC** | **Cases** | **Controls** | **p** | **AUROC** | **Cases** | **AUROC** | **Cases** | **AUROC** | **Cases** | **AUROC** | **Cases** | **AUROC** | **Cases** |
| **TRAFD1** | 0.67 (0.62 - 0.71) | 172 | 2,588 | 0.000 | 0.67 (0.6 - 0.74) | 68 | 0.67 (0.61 - 0.72) | 109 | 0.66 (0.61 - 0.7) | 207 | 0.67 (0.6 - 0.75) | 63 | 0.58 (0.48 - 0.68) | 35 |
| **GNG7** | 0.66 (0.61 - 0.72) | 138 | 3,244 | 0.002 | 0.67 (0.57 - 0.78) | 34 | 0.66 (0.58 - 0.73) | 75 | 0.65 (0.6 - 0.7) | 168 | 0.67 (0.6 - 0.74) | 63 | 0.59 (0.48 - 0.7) | 30 |
| **CD79B** | 0.66 (0.61 - 0.71) | 138 | 3,244 | 0.001 | 0.68 (0.58 - 0.78) | 34 | 0.69 (0.62 - 0.75) | 75 | 0.65 (0.61 - 0.7) | 168 | 0.64 (0.56 - 0.71) | 63 | 0.59 (0.48 - 0.7) | 30 |
| **RABL2A** | 0.66 (0.61 - 0.71) | 138 | 3,243 | 0.001 | 0.73 (0.64 - 0.83) | 34 | 0.69 (0.62 - 0.75) | 75 | 0.65 (0.61 - 0.7) | 168 | 0.64 (0.56 - 0.71) | 63 | 0.61 (0.5 - 0.73) | 30 |
| **BLK** | 0.65 (0.61 - 0.7) | 189 | 4,973 | 0.000 | 0.66 (0.59 - 0.73) | 73 | 0.67 (0.62 - 0.72) | 115 | 0.65 (0.6 - 0.69) | 232 | 0.63 (0.56 - 0.71) | 74 | 0.59 (0.49 - 0.7) | 43 |
| **WARS** | 0.65 (0.6 - 0.71) | 138 | 3,243 | 0.000 | 0.74 (0.64 - 0.83) | 34 | 0.66 (0.59 - 0.73) | 75 | 0.64 (0.59 - 0.69) | 168 | 0.65 (0.56 - 0.73) | 63 | 0.6 (0.5 - 0.7) | 30 |
| **ID3** | 0.65 (0.6 - 0.69) | 189 | 4,977 | 0.000 | 0.62 (0.55 - 0.69) | 73 | 0.64 (0.58 - 0.7) | 115 | 0.63 (0.59 - 0.67) | 232 | 0.66 (0.59 - 0.73) | 74 | 0.55 (0.44 - 0.65) | 43 |
| **GAS6** | 0.64 (0.59 - 0.68) | 178 | 4,628 | 0.000 | 0.71 (0.63 - 0.78) | 66 | 0.68 (0.62 - 0.73) | 107 | 0.64 (0.6 - 0.68) | 220 | 0.58 (0.5 - 0.66) | 71 | 0.63 (0.55 - 0.72) | 42 |
| **CCR6** | 0.64 (0.58 - 0.69) | 138 | 3,240 | 0.000 | 0.71 (0.61 - 0.81) | 34 | 0.68 (0.62 - 0.75) | 75 | 0.63 (0.59 - 0.68) | 168 | 0.58 (0.5 - 0.66) | 63 | 0.63 (0.51 - 0.75) | 30 |
| **RAB13** | 0.63 (0.58 - 0.68) | 138 | 3,244 | 0.000 | 0.66 (0.55 - 0.76) | 34 | 0.62 (0.55 - 0.69) | 75 | 0.65 (0.6 - 0.69) | 168 | 0.64 (0.57 - 0.71) | 63 | 0.72 (0.63 - 0.8) | 30 |
| **TUBGCP6** | 0.63 (0.58 - 0.67) | 189 | 4,982 | 0.000 | 0.63 (0.56 - 0.69) | 73 | 0.63 (0.58 - 0.69) | 115 | 0.62 (0.58 - 0.66) | 232 | 0.61 (0.54 - 0.68) | 74 | 0.59 (0.49 - 0.68) | 43 |
| **ZNF296** | 0.63 (0.57 - 0.68) | 138 | 3,216 | 0.000 | 0.72 (0.65 - 0.8) | 34 | 0.68 (0.61 - 0.74) | 75 | 0.62 (0.57 - 0.67) | 168 | 0.56 (0.48 - 0.64) | 63 | 0.58 (0.46 - 0.7) | 30 |
| **CD36** | 0.62 (0.57 - 0.67) | 138 | 3,242 | 0.000 | 0.68 (0.59 - 0.77) | 34 | 0.62 (0.56 - 0.69) | 75 | 0.62 (0.58 - 0.67) | 168 | 0.61 (0.53 - 0.7) | 63 | 0.64 (0.54 - 0.74) | 30 |
| **RAB33A** | 0.61 (0.56 - 0.67) | 138 | 3,234 | 0.000 | 0.64 (0.52 - 0.75) | 34 | 0.65 (0.58 - 0.72) | 75 | 0.6 (0.55 - 0.65) | 168 | 0.57 (0.49 - 0.65) | 63 | 0.55 (0.42 - 0.68) | 30 |
| **ZDHHC19** | 0.61 (0.56 - 0.66) | 137 | 3,189 | 0.000 | 0.62 (0.51 - 0.72) | 34 | 0.62 (0.55 - 0.68) | 75 | 0.59 (0.55 - 0.64) | 167 | 0.6 (0.52 - 0.68) | 62 | 0.51 (0.41 - 0.61) | 30 |
| **SDR39U1** | 0.61 (0.56 - 0.65) | 189 | 4,978 | 0.000 | 0.6 (0.53 - 0.67) | 73 | 0.61 (0.55 - 0.66) | 115 | 0.6 (0.56 - 0.64) | 232 | 0.6 (0.53 - 0.67) | 74 | 0.57 (0.48 - 0.67) | 43 |
| **CD1C** | 0.6 (0.56 - 0.65) | 189 | 4,974 | 0.000 | 0.64 (0.56 - 0.71) | 73 | 0.64 (0.58 - 0.7) | 115 | 0.58 (0.54 - 0.62) | 232 | 0.54 (0.48 - 0.61) | 74 | 0.49 (0.39 - 0.59) | 43 |

| | 0-12 months (primary analysis) | | | | 0-3 months | | 0-6 months | | 0-15 months | | 6-12 months | | 12-15 months | |
|---|---|---|---|---|---|---|---|---|---|---|---|---|---|---|
| **Signature** | **AUROC** | **Cases** | **Controls** | **p** | **AUROC** | **Cases** | **AUROC** | **Cases** | **AUROC** | **Cases** | **AUROC** | **Cases** | **AUROC** | **Cases** |
| **MAP7D3** | 0.59 (0.55 - 0.63) | 189 | 4,975 | 0.000 | 0.59 (0.52 - 0.66) | 73 | 0.59 (0.54 - 0.65) | 115 | 0.58 (0.54 - 0.62) | 232 | 0.58 (0.51 - 0.66) | 74 | 0.52 (0.42 - 0.62) | 43 |
| **CD3E** | 0.59 (0.54 - 0.65) | 138 | 3,244 | 0.000 | 0.67 (0.56 - 0.77) | 34 | 0.61 (0.54 - 0.69) | 75 | 0.59 (0.54 - 0.64) | 168 | 0.57 (0.49 - 0.65) | 63 | 0.56 (0.44 - 0.69) | 30 |
| **CD40LG** | 0.59 (0.54 - 0.65) | 138 | 3,244 | 0.000 | 0.61 (0.49 - 0.72) | 34 | 0.6 (0.52 - 0.68) | 75 | 0.57 (0.52 - 0.62) | 168 | 0.59 (0.51 - 0.67) | 63 | 0.52 (0.4 - 0.65) | 30 |
| **F2RL1** | 0.59 (0.54 - 0.64) | 137 | 3,100 | 0.000 | 0.62 (0.51 - 0.72) | 34 | 0.56 (0.48 - 0.63) | 75 | 0.57 (0.52 - 0.61) | 167 | 0.62 (0.56 - 0.69) | 62 | 0.52 (0.42 - 0.63) | 30 |
| **KLF2** | 0.59 (0.54 - 0.64) | 188 | 4,952 | 0.000 | 0.59 (0.51 - 0.66) | 72 | 0.58 (0.52 - 0.64) | 114 | 0.58 (0.54 - 0.63) | 231 | 0.6 (0.53 - 0.67) | 74 | 0.54 (0.45 - 0.64) | 43 |
| **S100A8** | 0.59 (0.54 - 0.64) | 138 | 3,244 | 0.000 | 0.68 (0.6 - 0.77) | 34 | 0.61 (0.54 - 0.68) | 75 | 0.59 (0.54 - 0.63) | 168 | 0.57 (0.49 - 0.64) | 63 | 0.58 (0.48 - 0.69) | 30 |
| **TRMT2A** | 0.59 (0.54 - 0.63) | 186 | 4,930 | 0.000 | 0.47 (0.4 - 0.55) | 72 | 0.57 (0.51 - 0.63) | 114 | 0.58 (0.54 - 0.62) | 229 | 0.61 (0.53 - 0.68) | 72 | 0.55 (0.46 - 0.64) | 43 |
| **KLHDC8B** | 0.58 (0.54 - 0.63) | 138 | 3,244 | 0.000 | 0.65 (0.56 - 0.74) | 34 | 0.65 (0.59 - 0.71) | 75 | 0.59 (0.54 - 0.63) | 168 | 0.49 (0.42 - 0.56) | 63 | 0.61 (0.49 - 0.72) | 30 |
| **NOD2** | 0.58 (0.53 - 0.64) | 138 | 3,244 | 0.000 | 0.59 (0.49 - 0.7) | 34 | 0.55 (0.48 - 0.63) | 75 | 0.58 (0.53 - 0.63) | 168 | 0.61 (0.54 - 0.69) | 63 | 0.56 (0.46 - 0.66) | 30 |
| **HK3** | 0.58 (0.53 - 0.63) | 138 | 3,244 | 0.000 | 0.65 (0.56 - 0.74) | 34 | 0.59 (0.53 - 0.66) | 75 | 0.57 (0.53 - 0.62) | 168 | 0.57 (0.49 - 0.65) | 63 | 0.54 (0.44 - 0.64) | 30 |
| **MAFB** | 0.58 (0.52 - 0.63) | 138 | 3,178 | 0.000 | 0.6 (0.49 - 0.71) | 34 | 0.55 (0.48 - 0.62) | 75 | 0.56 (0.52 - 0.61) | 168 | 0.61 (0.53 - 0.68) | 63 | 0.49 (0.38 - 0.6) | 30 |
| **KIF1B** | 0.57 (0.52 - 0.62) | 138 | 3,244 | 0.000 | 0.56 (0.46 - 0.66) | 34 | 0.56 (0.5 - 0.63) | 75 | 0.56 (0.52 - 0.61) | 168 | 0.59 (0.51 - 0.66) | 63 | 0.5 (0.39 - 0.6) | 30 |
| **BCL6** | 0.57 (0.51 - 0.62) | 138 | 3,244 | 0.000 | 0.62 (0.52 - 0.72) | 34 | 0.59 (0.52 - 0.66) | 75 | 0.56 (0.51 - 0.61) | 168 | 0.54 (0.46 - 0.61) | 63 | 0.54 (0.42 - 0.66) | 30 |
| **STT3A** | 0.55 (0.51 - 0.59) | 188 | 4,974 | 0.000 | 0.53 (0.46 - 0.6) | 73 | 0.54 (0.49 - 0.6) | 115 | 0.55 (0.51 - 0.58) | 231 | 0.56 (0.49 - 0.63) | 73 | 0.53 (0.44 - 0.62) | 43 |
| **LTF** | 0.55 (0.5 - 0.6) | 138 | 3,235 | 0.000 | 0.58 (0.48 - 0.68) | 34 | 0.57 (0.5 - 0.64) | 75 | 0.53 (0.49 - 0.57) | 168 | 0.53 (0.46 - 0.59) | 63 | 0.57 (0.47 - 0.66) | 30 |
| **PRDM1** | 0.55 (0.5 - 0.6) | 138 | 3,244 | 0.000 | 0.54 (0.43 - 0.64) | 34 | 0.52 (0.45 - 0.59) | 75 | 0.54 (0.49 - 0.58) | 168 | 0.59 (0.52 - 0.66) | 63 | 0.52 (0.41 - 0.62) | 30 |
| **LAG3** | 0.54 (0.49 - 0.6) | 138 | 3,244 | 0.000 | 0.62 (0.52 - 0.72) | 34 | 0.6 (0.53 - 0.67) | 75 | 0.55 (0.5 - 0.6) | 168 | 0.52 (0.44 - 0.61) | 63 | 0.6 (0.5 - 0.7) | 30 |

| | 0-12 months (primary analysis) | | | | 0-3 months | | 0-6 months | | 0-15 months | | 6-12 months | | 12-15 months | |
|---|---|---|---|---|---|---|---|---|---|---|---|---|---|---|
| Signature | AUROC | Cases | Controls | p | AUROC | Cases | AUROC | Cases | AUROC | Cases | AUROC | Cases | AUROC | Cases |
| **CDKN1C** | 0.54 (0.49 - 0.59) | 138 | 3,242 | 0.000 | 0.52 (0.44 - 0.61) | 34 | 0.57 (0.51 - 0.64) | 75 | 0.55 (0.51 - 0.6) | 168 | 0.49 (0.42 - 0.56) | 63 | 0.61 (0.52 - 0.71) | 30 |
| **SLPI** | 0.54 (0.49 - 0.59) | 138 | 3,244 | 0.000 | 0.65 (0.56 - 0.74) | 34 | 0.56 (0.5 - 0.63) | 75 | 0.54 (0.49 - 0.59) | 168 | 0.49 (0.42 - 0.57) | 63 | 0.55 (0.45 - 0.66) | 30 |
| **TNIP1** | 0.54 (0.49 - 0.59) | 138 | 3,244 | 0.000 | 0.5 (0.4 - 0.61) | 34 | 0.53 (0.46 - 0.6) | 75 | 0.53 (0.49 - 0.58) | 168 | 0.55 (0.48 - 0.62) | 63 | 0.49 (0.39 - 0.59) | 30 |
| **MMP9** | 0.54 (0.49 - 0.58) | 138 | 3,244 | 0.000 | 0.47 (0.38 - 0.57) | 34 | 0.56 (0.5 - 0.63) | 75 | 0.53 (0.48 - 0.57) | 168 | 0.5 (0.43 - 0.57) | 63 | 0.51 (0.4 - 0.63) | 30 |
| **CD160** | 0.54 (0.48 - 0.59) | 138 | 3,243 | 0.000 | 0.48 (0.37 - 0.59) | 34 | 0.56 (0.49 - 0.63) | 75 | 0.54 (0.49 - 0.59) | 168 | 0.49 (0.42 - 0.57) | 63 | 0.55 (0.43 - 0.66) | 30 |
| **CD74** | 0.53 (0.48 - 0.59) | 138 | 3,244 | 0.000 | 0.49 (0.37 - 0.6) | 34 | 0.51 (0.44 - 0.58) | 75 | 0.53 (0.48 - 0.58) | 168 | 0.56 (0.48 - 0.65) | 63 | 0.51 (0.4 - 0.62) | 30 |
| **RBBP8** | 0.53 (0.48 - 0.58) | 138 | 3,235 | 0.000 | 0.57 (0.46 - 0.68) | 34 | 0.51 (0.44 - 0.58) | 75 | 0.53 (0.48 - 0.58) | 168 | 0.58 (0.51 - 0.65) | 63 | 0.49 (0.37 - 0.6) | 30 |
| **C5** | 0.52 (0.47 - 0.58) | 138 | 3,187 | 0.000 | 0.53 (0.43 - 0.62) | 34 | 0.5 (0.43 - 0.57) | 75 | 0.48 (0.43 - 0.53) | 167 | 0.55 (0.47 - 0.63) | 63 | 0.51 (0.4 - 0.62) | 29 |
| **MPO** | 0.52 (0.47 - 0.57) | 138 | 3,234 | 0.000 | 0.49 (0.39 - 0.58) | 34 | 0.49 (0.43 - 0.55) | 75 | 0.53 (0.48 - 0.57) | 168 | 0.53 (0.46 - 0.61) | 63 | 0.44 (0.34 - 0.54) | 30 |
| **UCP2** | 0.52 (0.47 - 0.56) | 189 | 4,982 | 0.000 | 0.49 (0.42 - 0.56) | 73 | 0.52 (0.46 - 0.58) | 115 | 0.49 (0.45 - 0.53) | 232 | 0.49 (0.41 - 0.57) | 74 | 0.51 (0.41 - 0.61) | 43 |
| **MTRF1L** | 0.51 (0.46 - 0.56) | 138 | 3,244 | 0.000 | 0.51 (0.41 - 0.62) | 34 | 0.51 (0.44 - 0.58) | 75 | 0.51 (0.47 - 0.56) | 168 | 0.54 (0.47 - 0.62) | 63 | 0.51 (0.41 - 0.61) | 30 |
| **CYP4F3** | 0.5 (0.45 - 0.55) | 138 | 3,237 | 0.000 | 0.53 (0.43 - 0.63) | 34 | 0.49 (0.42 - 0.55) | 75 | 0.5 (0.45 - 0.54) | 168 | 0.51 (0.44 - 0.58) | 63 | 0.49 (0.37 - 0.6) | 30 |
| **ACTA2** | 0.5 (0.44 - 0.55) | 138 | 3,243 | 0.000 | 0.54 (0.43 - 0.65) | 34 | 0.52 (0.45 - 0.59) | 75 | 0.51 (0.46 - 0.56) | 168 | 0.54 (0.47 - 0.61) | 63 | 0.57 (0.46 - 0.68) | 30 |
| **KAZN** | 0.49 (0.44 - 0.55) | 138 | 3,218 | 0.000 | 0.52 (0.42 - 0.63) | 34 | 0.53 (0.46 - 0.59) | 75 | 0.51 (0.46 - 0.55) | 168 | 0.52 (0.45 - 0.6) | 63 | 0.55 (0.44 - 0.66) | 30 |
| **GZMA** | 0.49 (0.44 - 0.54) | 138 | 3,244 | 0.000 | 0.52 (0.41 - 0.64) | 34 | 0.56 (0.49 - 0.63) | 75 | 0.5 (0.45 - 0.54) | 168 | 0.55 (0.48 - 0.63) | 63 | 0.52 (0.41 - 0.63) | 30 |
| **GNGT1** | 0.49 (0.43 - 0.55) | 138 | 3,243 | 0.000 | 0.58 (0.47 - 0.7) | 34 | 0.5 (0.42 - 0.58) | 75 | 0.49 (0.43 - 0.54) | 168 | 0.52 (0.44 - 0.61) | 63 | 0.53 (0.41 - 0.65) | 30 |
| **TMCC1** | 0.47 (0.42 - 0.53) | 138 | 3,244 | 0.000 | 0.59 (0.48 - 0.69) | 34 | 0.47 (0.39 - 0.54) | 75 | 0.49 (0.44 - 0.54) | 168 | 0.48 (0.41 - 0.56) | 63 | 0.56 (0.45 - 0.68) | 30 |

***Supplementary table 3: Performance metrics table of all multi-gene signatures (subclinical TB 0-12 months)***

Performance metrics of all multi-gene signatures to discriminate subclinical TB cases from non-progressors at an interval from 12 months from sampling to disease are shown in this table. Equivalence to Roe3, the best multi-gene signature, was defined as an adjusted p value >0.05 in pairwise Delong test. Performance metrics include estimates and 95% confidence intervals for the Area Under the Receiver Operating Curve (AUROC) as well as sensitivity and specificity at the maximum Youden index calculated from the one-stage meta-analysis.

| Signature | AUROC | Sensitivity | Specificity | N | Cases | Controls | p |
|---|---|---|---|---|---|---|---|
| **Roe3** | 0.77 (0.73 - 0.81) | 0.74 (0.67 - 0.79) | 0.7 (0.69 - 0.71) | 5,171 | 189 | 4,982 | Ref |
| **Maertzdorf4** | 0.74 (0.7 - 0.78) | 0.65 (0.58 - 0.72) | 0.73 (0.72 - 0.74) | 5,062 | 189 | 4,873 | 0.032 |
| **Suliman4** | 0.74 (0.69 - 0.8) | 0.64 (0.55 - 0.72) | 0.76 (0.74 - 0.77) | 4,451 | 111 | 4,340 | 0.001 |
| **PennNicholson6** | 0.74 (0.69 - 0.78) | 0.62 (0.54 - 0.7) | 0.76 (0.75 - 0.77) | 4,901 | 143 | 4,758 | 0.478 |
| **Francisco2** | 0.73 (0.69 - 0.77) | 0.63 (0.56 - 0.7) | 0.74 (0.73 - 0.75) | 5,140 | 188 | 4,952 | 0.012 |
| **Darboe11** | 0.73 (0.68 - 0.78) | 0.77 (0.69 - 0.83) | 0.6 (0.58 - 0.62) | 2,479 | 129 | 2,350 | 0.103 |
| **Sweeney3** | 0.72 (0.68 - 0.77) | 0.49 (0.42 - 0.56) | 0.88 (0.87 - 0.89) | 5,140 | 188 | 4,952 | 0.010 |
| **Thompson5** | 0.67 (0.63 - 0.72) | 0.61 (0.54 - 0.68) | 0.65 (0.63 - 0.66) | 5,156 | 188 | 4,968 | 0.000 |

***Supplementary figure 10: Forest plot of AUROC estimates for best performing single-gene transcripts (subclinical TB 0-12 months)***

Study-level and pooled AUROC estimates (95% CI) for the best performing single-gene transcripts to discriminate subclinical TB cases from non-progressors at an interval from 12 months from sampling to disease are shown in this forest plot.

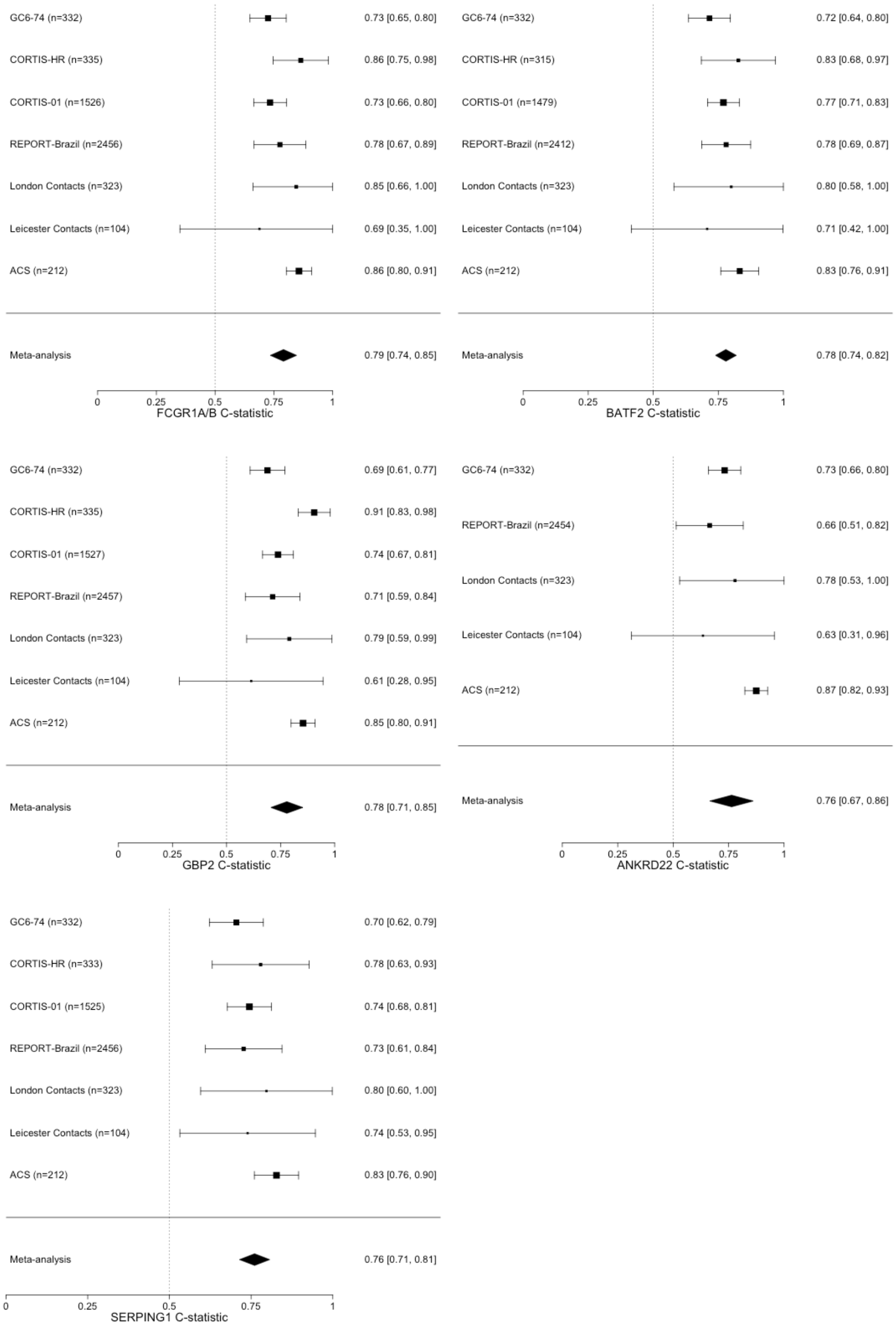

***Supplementary table 4: Performance metrics table of BATF2, IGRA and a combined approach (subclinical TB 0-12 months)***

Performance metrics are shown for subclinical TB (0-12 months) of BATF2 (threshold set at maximum Youden index), IGRA (threshold set at standard cutoff of 0.35 IU/ml) and a combined approach of BATF2 and IGRA, stratified by setting. Performance metrics include sensitivity (95% CIs), specificity (95% CIs), positive predictive value (PPV [95% CIs]), negative predictive value (NPV [95% CIs]), false positive rate (FPR), false negative rate (FNR), true positive rate (TPR), true negative rate (TNR). A prior probability of 1% was used.

| Test | Setting | Sensitivity (95% CI) | Specificity (95% CI) | PPV (95% CI) | NPV (95% CI) | FPR | FNR | TPR | TNR | Cases | Controls |
|---|---|---|---|---|---|---|---|---|---|---|---|
| **IGRA** | All | 0.87 (0.79 - 0.92) | 0.54 (0.53 - 0.56) | 0.019 (0.017 - 0.021) | 0.998 (0.996 - 0.999) | 0.456 | 0.130 | 0.870 | 0.544 | 92 | 3,243 |
| **IGRA** | High burden | 0.87 (0.77 - 0.93) | 0.32 (0.3 - 0.35) | 0.013 (0.011 - 0.014) | 0.996 (0.992 - 0.998) | 0.677 | 0.132 | 0.868 | 0.323 | 68 | 1,521 |
| **IGRA** | Low burden | 0.88 (0.69 - 0.96) | 0.74 (0.72 - 0.76) | 0.033 (0.024 - 0.039) | 0.998 (0.996 - 0.999) | 0.260 | 0.125 | 0.875 | 0.740 | 24 | 1,722 |
| **BATF2** | All | 0.75 (0.65 - 0.83) | 0.69 (0.68 - 0.71) | 0.024 (0.02 - 0.028) | 0.996 (0.995 - 0.998) | 0.306 | 0.250 | 0.750 | 0.694 | 92 | 3,243 |
| **BATF2** | High burden | 0.78 (0.67 - 0.86) | 0.67 (0.64 - 0.69) | 0.023 (0.018 - 0.027) | 0.997 (0.995 - 0.998) | 0.332 | 0.221 | 0.779 | 0.668 | 68 | 1,521 |
| **BATF2** | Low burden | 0.67 (0.47 - 0.82) | 0.72 (0.7 - 0.74) | 0.024 (0.016 - 0.031) | 0.995 (0.992 - 0.998) | 0.282 | 0.333 | 0.667 | 0.718 | 24 | 1,722 |
| **IGRA & BATF2** | All | 0.64 (0.54 - 0.73) | 0.85 (0.84 - 0.86) | 0.041 (0.033 - 0.05) | 0.996 (0.994 - 0.997) | 0.150 | 0.359 | 0.641 | 0.850 | 92 | 3,243 |
| **IGRA & BATF2** | High burden | 0.66 (0.54 - 0.76) | 0.77 (0.75 - 0.79) | 0.028 (0.021 - 0.035) | 0.996 (0.994 - 0.997) | 0.226 | 0.338 | 0.662 | 0.774 | 68 | 1,521 |
| **IGRA & BATF2** | Low burden | 0.58 (0.39 - 0.76) | 0.92 (0.9 - 0.93) | 0.068 (0.038 - 0.099) | 0.995 (0.993 - 0.997) | 0.084 | 0.417 | 0.583 | 0.916 | 24 | 1,722 |

***Supplementary figure 11: Diagnostic performance for subclinical TB (0-6 months) of BATF2, IGRA and a combined approach shown in receiver operating space, by setting***

This plot compares the diagnostic performance for subclinical TB over a 0-6 month interval of BATF2 (threshold set at maximum Youden Index), IGRA (threshold set at standard cutoff of 0.35 IU/ml) and a combined approach of BATF2 and IGRA in the receiver operating space with sensitivity on the y axis and 1-specificity on the x axis, stratified by setting. Shown as point estimates with 95% confidence intervals (boxes). Dotted lines represent the 75% WHO Target Product Profile minimum sensitivity and specificity for a TB progression test. Dashed lines represent positive predictive values (PPVs) of 1%, 3% and 5%, based on a 1% prior probability.

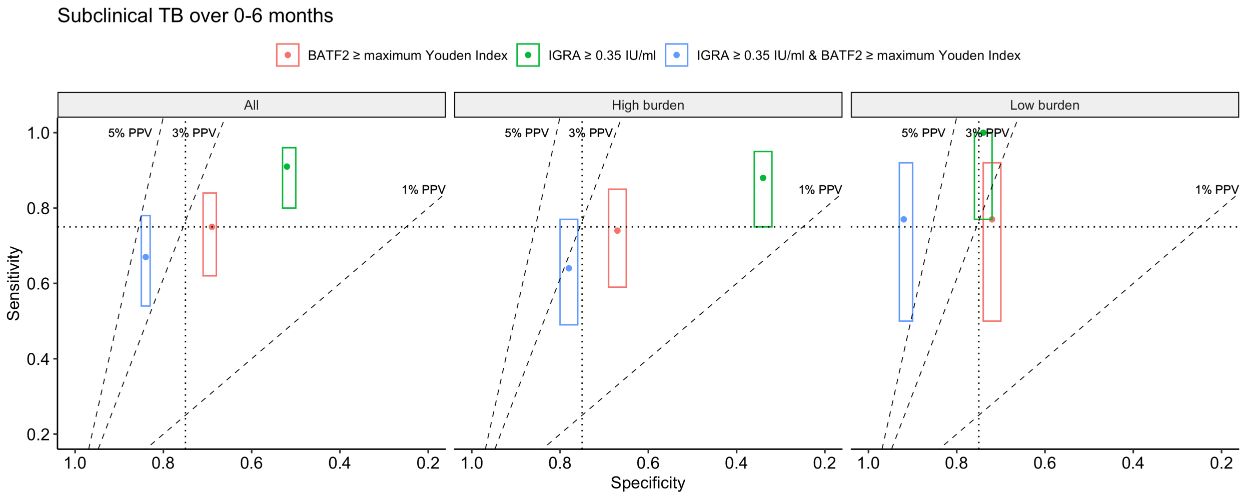

***Supplementary figure 12: Diagnostic performance for subclinical TB of BATF2, IGRA, and a combined approach shown in receiver operating space and in a decision curve analysis, by setting, using a 2% prior probability***

Supplementary figure 12A compares the diagnostic performance for subclinical TB over a 0-12 month interval of BATF2 (threshold set at maximum Youden Index), IGRA (threshold set at standard cutoff of 0.35 IU/ml) and a combined approach of BATF2 and IGRA in the receiver operating space with sensitivity on the y axis and 1-specificity on the x axis, stratified by setting. Only participants with results for both tests were included in this analysis. Shown as point estimates with 95% confidence intervals (boxes). Dotted lines represent the 75% WHO minimum Target Product Profile sensitivity and specificity for a TB progression test. Dashed lines represent positive predictive values (PPVs) of 3%, 5% and 10%, based on a 2% prior probability.

Supplementary figure 12B is a decision curve analysis where each test is compared to default strategies of treating all or treating no persons, stratified by setting. Threshold probability is the risk of TB disease at which a clinician or patient would opt for preventative therapy and is the reciprocal of the number-willing-to-treat to prevent a single case. Net benefit is calculated at a range of threshold probabilities as the true positive rate minus a weighted false positive rate, where the weighting is the threshold probability. Since the contributing datasets included case-control analyses, the cumulative TB risk was fixed at 2%.

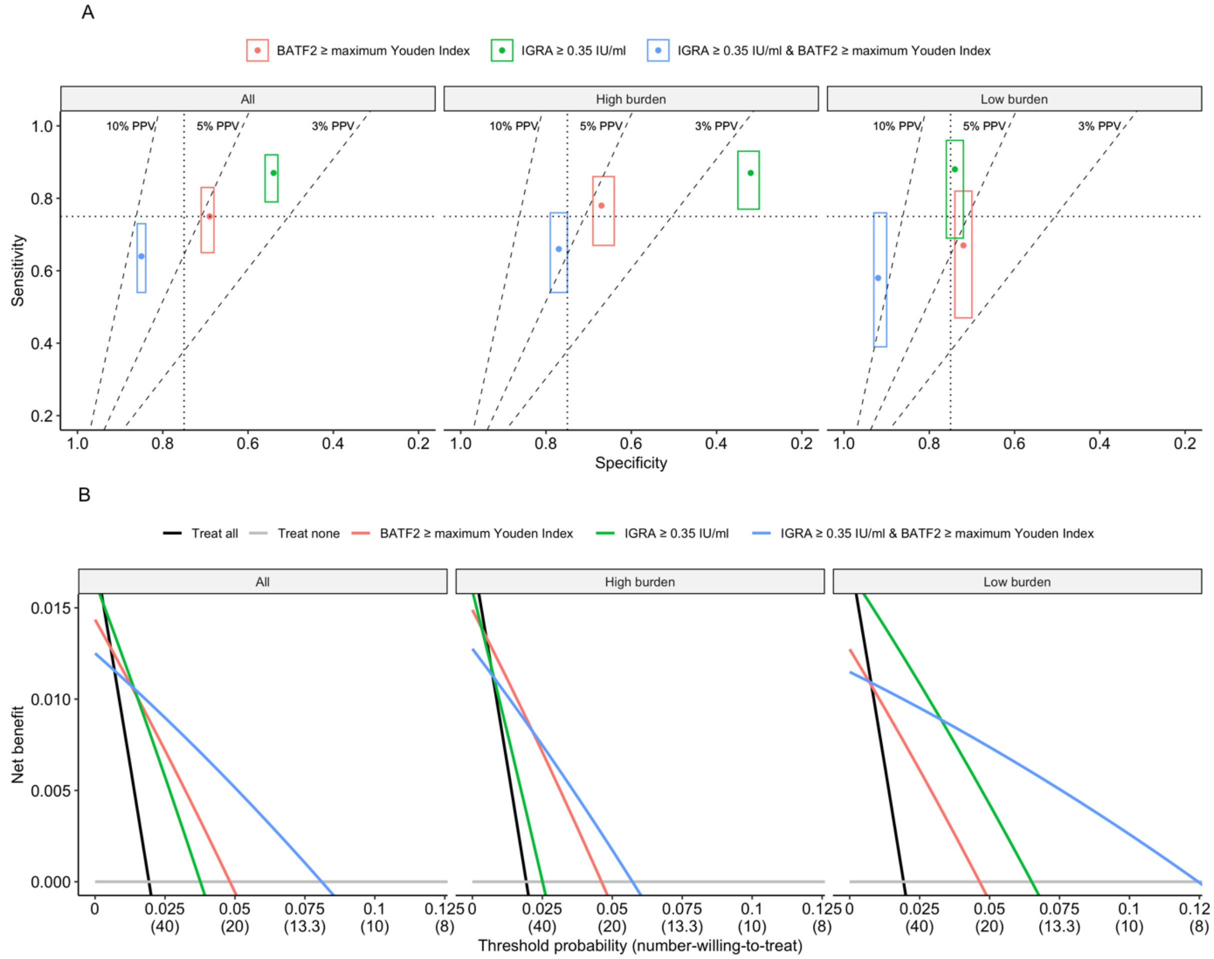

***Supplementary table 5: Table showing AUROC estimates (95% CI) from sensitivity analyses of best performing multi-gene signature and equivalent single-gene transcripts***

The primary analysis is of diagnostic performance for subclinical TB (0 - 12 months) from a one-stage meta-analysis, excluding recipients of TB Preventative Therapy (TPT) and considering serial samples from a single individual as independent samples. Secondary analyses included a two-stage meta-analysis, an analysis including only one randomly-selected sample per individual and an analysis including recipients of TPT.

| Signature | Primary analysis | Two-stage meta-analysis | Single sample per individual | Including TPT recipients |
|---|---|---|---|---|
| **BATF2** | 0.77 (0.73 - 0.81) | 0.78 (0.74 - 0.82) | 0.75 (0.71 - 0.8) | 0.77 (0.73 - 0.8) |
| **FCGR1A/B** | 0.77 (0.73 - 0.81) | 0.79 (0.74 - 0.85) | 0.75 (0.71 - 0.79) | 0.77 (0.73 - 0.8) |
| **Roe3** | 0.77 (0.73 - 0.81) | 0.8 (0.74 - 0.86) | 0.76 (0.72 - 0.8) | 0.77 (0.73 - 0.81) |
| **ANKRD22** | 0.77 (0.72 - 0.81) | 0.76 (0.67 - 0.86) | 0.74 (0.68 - 0.8) | 0.76 (0.72 - 0.81) |
| **GBP2** | 0.75 (0.71 - 0.79) | 0.78 (0.71 - 0.85) | 0.74 (0.7 - 0.78) | 0.75 (0.71 - 0.79) |
| **SERPING1** | 0.75 (0.71 - 0.79) | 0.76 (0.71 - 0.81) | 0.74 (0.69 - 0.78) | 0.75 (0.71 - 0.79) |

***Supplementary table 6: Table showing AUROC estimates (95% CI) of multi-gene signatures that were derived from datasets used in analysis (subclinical TB 0-12 months)***

Three multi-gene signatures were originally derived from datasets including in this analysis; PennNicholson6 and Darboe 11 from ACS, Suliman4 from GC6-74. In the primary analysis these signatures were not calculated for samples from these derivation datasets, however in a secondary analysis they were calculated for samples from these datasets.

| Signature | Primary analysis | Including derivation dataset |
|---|---|---|
| **Suliman4** | 0.74 (0.69 - 0.8) | 0.74 (0.7 - 0.78) |
| **Penn-Nicholson6** | 0.74 (0.69 - 0.78) | 0.77 (0.73 - 0.81) |
| **Darboe11** | 0.73 (0.68 - 0.78) | 0.76 (0.72 - 0.8) |
